# Cohort-scale Spatial Host-Microbiome Predicts Post-Resection Recurrence in Colorectal Cancer

**DOI:** 10.64898/2026.09.29.26363734

**Authors:** Feng Guo, Hailiang Sun, Chenxi Hu, Minsheng Hao, Youyang Wan, Chuxiao Xiong, Feng Gao, Lung-Yi Mak, Xin Deng, Ajay Goel, Jia Ke, Jianzhu Ma, Peng Shi

## Abstract

The tumor microenvironment in colorectal cancer (CRC) is a heterogeneous ecosystem in which host cells and microbial communities interact dynamically, influencing disease progression. However, the clinical utility is limited by the lack of a scalable spatial host-microbiome technique and by insufficient integration of artificial intelligence for the interpretation of high-dimensional multi-omics data. To overcome the barriers, we present AlphaFISH, a platform technology integrating both technical and computational innovations for multi-omics spatial analysis of clinical biopsies at subcellular resolution. The system uses a sequencing-free, high-throughput, spatial profiling technique to construct, to date, the largest clinical spatial transcriptomics and spatial microbiome datasets acquired from 149 colorectal biopsies from 68 human subjects, supported by a comprehensive scRNA-seq atlas covering 4.27 million cells across 650 patients for robust cell annotation. Deep learning of the cohort-scale dual-omics data, consisting of more than 10 million subcellular sampling vectors, enables the development of a transformer model with joint embeddings of gene expression, spatial architecture, and the microbial microenvironment in colon tissues, achieving nearly 90% accuracy in predicting CRC-associated pathological features using unseen spatial omics inputs. The AI interrogation further predicts tumour recurrence at 81% accuracy in 28 patients followed within 1 year post tumor resection period. AlphaFISH reveals that spatial interactions between *Fusobacterium* and cellular niche consisting of tumor and T cells serve as key markers of CRC malignancy, progression, and recurrence, indicating the critical role of spatial bacterial-immune crosstalks.

## Introduction

The tumor microenvironment in colorectal cancer (CRC) constitutes a complex ecosystem where host cells and microbial communities are engaged in dynamic and spatially heterogeneous crosstalk that drives disease progression^1^ that drives disease progression and tumor recurrence. Bulk omics approaches have established substantial correlations between specific microbes and transcriptional programs^2^. For example, *Fusobacterium nucleatum* was reported to be associated with β-catenin-mediated cell proliferation^3^. However, most of these studies lack the spatial context to resolve whether host-microbial interactions^4,5^, which could be deciphered via emerging spatial profiling technologies to provide precise co-localization of bacterial and host transcriptomic information^6^. A recent seminal work in mice revealed that specific bacterial genera, like *Pseudobutyrivibri and Intestimonasco*, respectively colocalize with colonocytes or neurons expressing different microbial receptors; immune cells expressing microbial response genes directly link particular microbes to functional host responses in the crypt niches^7^.

Despite the potential, current spatial profiling techniques struggle to simultaneously capture host transcriptomics and microbial signatures^8–10^. Most sequencing-based platforms use a poly(T) probe to target polyadenylated eukaryotic mRNAs, and simultaneously use 16S or 18S priming probes for microbial RNAs ^7,11^. However, the robust cell walls of many microbes hinder RNA accessibility; microbial RNAs are also often low in abundance and difficult to distinguish from the dominant host RNAs, leading to low detection sensitivity and limited taxonomic resolution. Spatial transcriptomics platforms typically lack the fine spatial resolution needed to resolve microbial micro-niches. In many cases, the requirement of tissue processing can degrade or displace microbial RNA, further disrupting spatial accuracy^12^. These challenges highlight the need for scalable dual-omics spatial techniques that simultaneously detect host and microbial transcriptomes in situ. Accordingly, technical innovation to develop low-cost, high-throughput spatial host-microbiome techniques is critical. Equally important is the generation of large, clinically relevant spatial omics datasets to fuel data interpretation challenges inherent in these complex datasets.

Particularly, artificial intelligence (AI) is playing an increasingly transformative role in the field of spatial omics, which enables the mapping of gene expression of cellular architecture within large tissues^13,14^. Some efforts have been made to explore the integration of multi-omics data for understanding CRC mechanisms in tissue-specific contexts^15,16^. The integration of AI, such as deep learning models like AlphaFold ^17,18^, offers promise for enhancing the spatial resolution and interpretability of molecular interactions in complex biological samples. AlphaFold’s ability to predict three-dimensional spatial interactions among amino acid motifs suggests a transferable capability for spatial omics to resolve intricate tissue microenvironments at the molecular level. However, significant challenges remain in applying AI effectively to predict clinical trajectories, such as postoperative progression from clinically derived spatial omics data. One of the primary limitations is the lack of reliable, large-scale spatial omics datasets from human disease samples. This is largely due to the prohibitive cost and technical complexity associated with existing spatial omics platforms, which hinder accessibility and scalability in routine clinical workflows ^19,20^. The diversity of technical solutions and proprietary platforms leads to a lack of standardization in data formats, complicating cross-study comparisons and the development of universal AI models^21,22^.

To address the pressing challenges, especially the need for scalable, clinically viable technologies and comprehensive clinical datasets to adopt spatial host-microbiome in CRC, we introduce a suite of technical and analytical innovations. First, we develop a sequencing-free, high-throughput spatial host-microbiome platform specifically designed for acute clinical biopsies. Central to this platform is a nanoprobe biochip that enables rapid, multiplexed isolation of 41 host transcripts and 51 genus-specific bacterial RNAs from acute CRC tissue biopsies, achieving single-cell spatial resolution to allow for precise, simultaneous profiling of host cell and microbial gene expression directly from patient-derived samples. To overcome data limitations, we have constructed, to date, the largest clinical spatial multi-omics data library, comprising 149 colorectal biopsies from 68 human subjects, including 51 CRC patients, 15 Crohn’s disease (CD) patients, and 2 healthy individuals. This dataset captures diverse spatial phenotypes across disease states and serves as a foundation for machine learning. Building on this, we developed AlphaFISH, a transformer-based deep learning model adapted from AlphaFold’s architecture. AlphaFISH learns joint spatial embeddings that integrate gene expression, spatial context, and microbial microenvironment. It demonstrates nearly 90% accuracy in predicting multiple CRC-related pathological features in unseen spatial host-microbiome inputs. Beyond diagnostic classification, it further predict post-resection secondary tumor recurrence with 81% accuracy in a one year longitudinal cohort of 28 CRC patients. Beyond pathological prediction, AlphaFISH reveals novel biological insights into CRC pathogenesis, identifying NK cell activation as a signature associated with tumor malignancy, uncovering spatial reorganization of key bacterial genera, including the well-documented interaction between Fusobacterium and tumor cells. It also highlights a previously underappreciated interaction between Haemophilus and T cells within the tumor immune niche. These results suggest the critical role of spatial bacterial-immune crosstalks in CRC malignancy, progression, and recurrence.

## Results

### System design

The AlphaFISH system relies on both technical and computational innovations (**Figure 1**). The technical foundation builds on the continuous development of a biochip-based, sequencing-free, high-throughput spatial multi-omics platform tailored for large-scale, acute clinical analysis ^23–25^. At its core, the biochip integrates nanoprobe technology that rapidly isolates 41 host transcripts and 51 genus-specific bacterial RNAs directly from clinical biopsy tissues. This enables spatially resolved profiling of host and microbiome interactions at single-cell resolution, without the need for conventional sequencing workflows^26^. The platform is optimized for clinical scalability, allowing for rapid processing of multiple samples. Using this technique, the largest spatial multi-omics clinical dataset to date was generated, comprising 149 colorectal biopsy samples from 68 human subjects, including 51 CRC patients, 15 Crohn’s disease patients, and 2 healthy controls. To analyze this complex data, a transformer-based deep learning model was computationally developed. AlphaFISH learns joint embeddings that integrate spatial context, gene expression, and microbial microenvironment, enabling it to accurately predict CRC-related pathological features from previously unseen spatial host-microbiome inputs. Moreover, AlphaFISH uncovers spatial phenotypes associated with immune cell activation and microbial reorganization across CRC progression stages, providing a robust computational framework for decoding the spatial complexity of host-microbiome interactions in disease.

**Figure 1.**
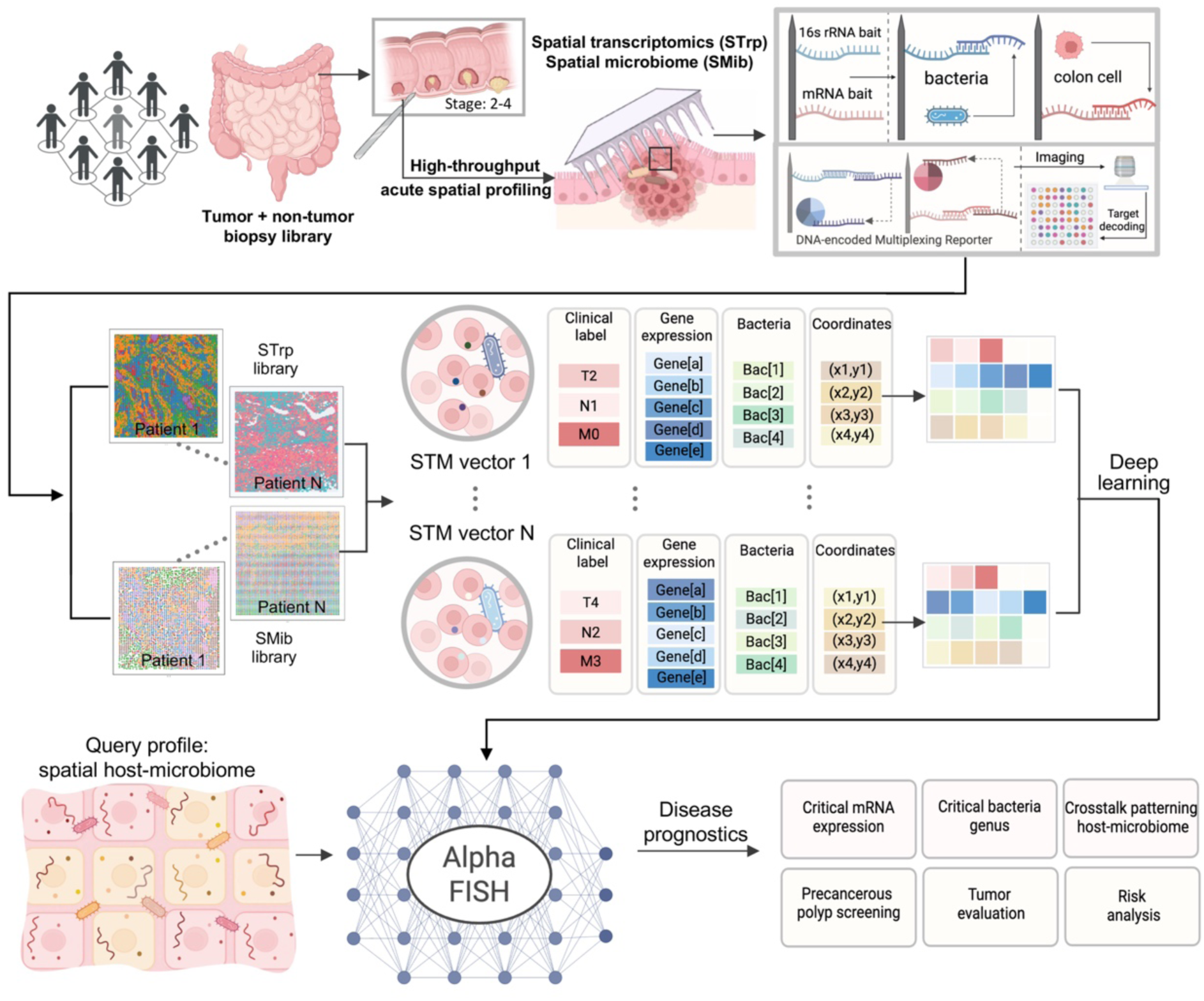
Schematic illustration of AlphaFISH: transformer model for learning and explaining high-throughput spatial host-microbiome profiling of clinical biopsy tissues. The technical innovation relies on a biochip-based, sequencing-free, high-throughput spatial multi-omics platform, developed for large-scale, high-throughput analysis of clinical samples. The nanostructured biochip enables rapid isolation of 41 host transcripts and 51 genus-specific bacterial RNAs for spatially resolved host-microbiome profiling at single-cell resolution. The largest clinical spatial multi-omics data library, encompassing both transcriptomics and microbiome data, is constructed from 149 colorectal biopsies from 68 human subjects, including 51 CRC patients, 15 Crohn’s disease patients, and 2 healthy subjects. A transformer-based model, AlphaFISH, is computationally developed by learning joint embeddings of spatial context, gene expression, and microbial microenviroment, predicting multiple aspects of CRC related pathologies in unseen spatial host-microbiome inputs, and further deciphering complex host–microbiome spatial phenotypes in the pathogenesis and disease progress of CRC, emphasizing the critical role of immune cells activation and interactive microbial reorganization at different stage of CRC.

### Dual-omics spatial profiling of host mRNAs and bacterial rRNA in acute biopsy tissues

To enable dual-omics spatial profiling of host transcriptomes and microbial communities in human clinical biopsy samples, the technical core of AlphaFISH relies on a sequencing-free, high-resolution, imaging-based spatial omics platform. The system utilizes a nanostructured biochip (1×1 mm²) containing ∼60,000 nanoprobes with a 0.8 µm diameter and a 4 µm pitch (**Suppl. Figure S1**), each functionalized with polyT and 16S rRNA-targeting bait sequences (**Figure 2a**). These dual baits enable parallel extraction of poly(A)-tailed mRNAs and bacterial 16S rRNAs from an interfacing tissue, respectively. The captured targets are hybridized to spectrally encoded DNA reporters and decoded via 7-channel confocal microscopy (**Suppl. Figure S2**), enabling the generation of high-dimensional molecular maps with single-cell spatial resolution (**Figure 2a**).

**Figure 2.**
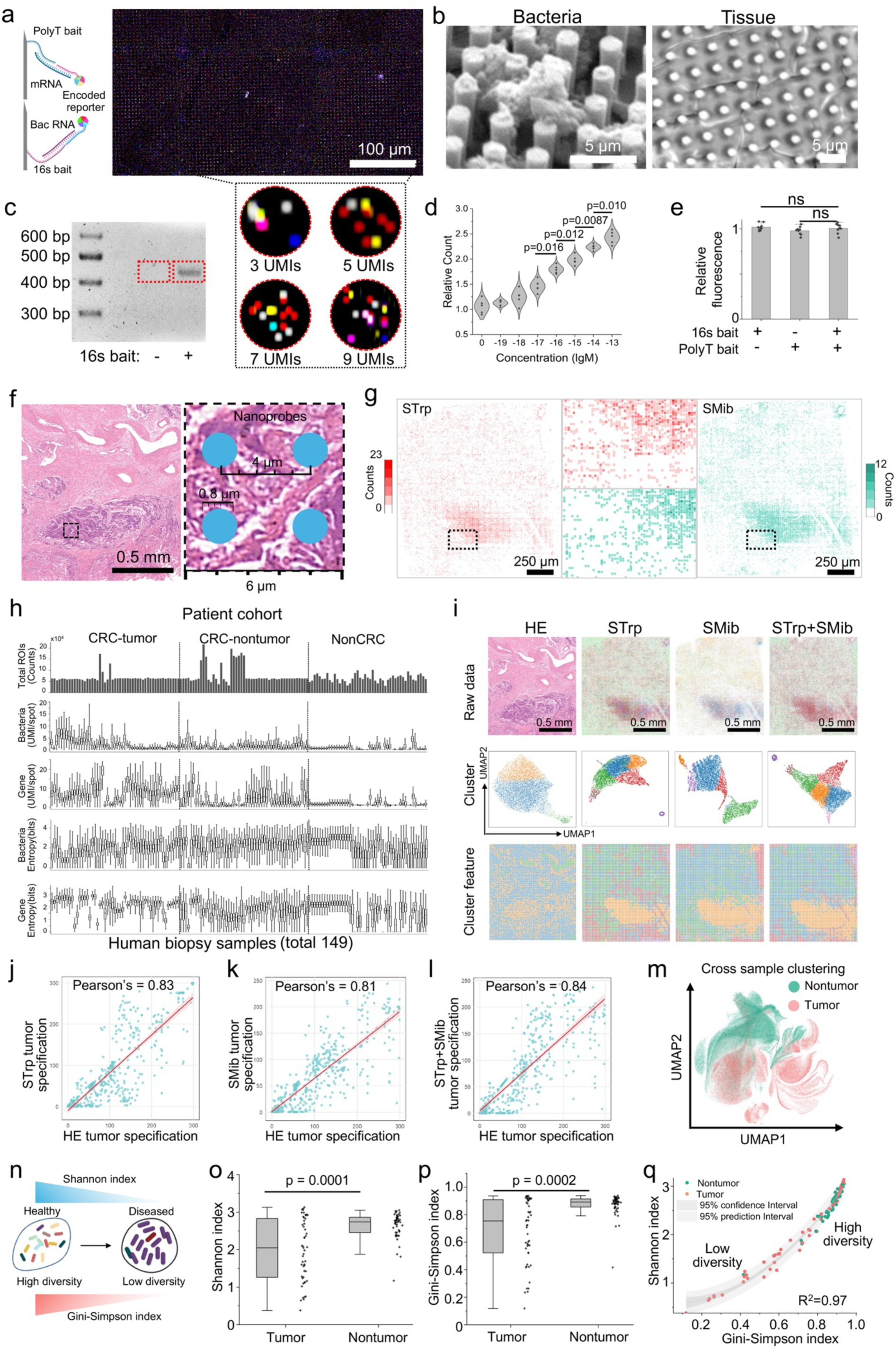
Dual-omics spatial profiling of host mRNAs and bacterial rRNA in acute biopsy tissues. **(a)** Schematic illustrating the technical details of AlphaFISH: a nanostructured biochip that enables imaging-based spatial multi-omics, resolving both transcriptomics and the microbiome. The nanoprobes are functionalized polyT and 16S-priming sequences, working with the specific baits to extract poly(A)-tailed mRNA via bacterial 16S rRNA, respectively. A 1×1 mm^2^ biochip contains ^∼^6×10^4 nanoprobes, each serving as the basic unit to achieve a spatial resolution of 4 µm. The captured targets hybridize with spectrum-encoded DNA reporters, which are decoded by 7-channel confocal imaging. Enlarged view showing 13 unique molecular identifiers (UMIs) decoded around an individual nanoprobe. Scale bar, 4 µm. **(b)** SEM images showing the interface of the nanoprobe array on a biochip with bacteria (left) and tissue (right). Scale bars, 4 µm. **(c)** Validation of nanoprobe-assisted extraction of bacterial RNAs. Nanoprobes with or without 16S-priming sequences are interfaced with bacteria (E.coli). Successful extraction of bacterial RNAs is confirmed by reverse-transcription and amplification (RT-PCR) and electrophoresis, giving a specific band at 400-500 bp, targeting a key phylogenetic marker of the V3–V4 hypervariable region. **(d)** A violin plot shows the sensitivity assessment of bacterial rRNA detection using synthetic RNA fragments titrated from 10^−13^M to 10^−19^M. Dots represent replicates, n=5; a two-tailed t-test calculates p-values. **(e)** Assessment of competitive extraction of mRNAs and bacterial RNAs using a biochip with dual-bait functionalization of polyT and 16S priming sequences. Three conditions are tested: polyT only, 16S priming sequence only, or dual-bait (polyT+16S priming sequences, 1:1 molar ratio); the total mole concentration of the bait sequences is kept constant, analyzed by two-tailed t-test, n=9, ns: not significant. Data are presented as the mean±SD. **(f)** H&E-stained section of a human CRC biopsy examined by a biochip. Scale bar, 0.5 mm. The enlarged view shows a schematic illustration of the tissue-nanoprobe (blue dots) interface. The biochip is fabricated as an array of nanoprobes (0.8 µm diameter, 4 µm pitch). **(g)** Representative results of dual omics spatial profiling in **(f)**. The UMI abundance of spatial transcriptomics (STrp, red, left) and spatial microbiome (SMib, cyan, right). The boxed region is enlarged in the center column. Scale bar, 250 µm. **(h)** Data quality metrics across 149 human colon biopsy samples (51 CRC-tumor, 51 CRC-nontumor, 47 non-CRC) from 68 patients. Top: Total counts of ROIs (signal positive nanoprobes) for each sample (^∼^60,000 for 1×1 mm^2^ biochip; ^∼^160,000 for 1.6×1.6 mm^2^ biochip). 2^nd^ & 3^rd^ row: UMI counts per ROI for different bacterial genus and gene transcripts. 4^th^ & 5^th^ row: entropy of detected bacteria and genes of all ROIs across each biochip. These metrics confirm robust and reliable dual-omics profiling. **(i)** Concordance between AlphaFISH-derived spatial omics profiles and H&E morphology features. Top: Raw H&E image and corresponding molecular density maps (STrp, SMib, STrp+SMib). Middle: UMAP showing clustering features based on individual-sample analysis. Bottom: Spatial distribution of specific clustering features. H&E features are extracted via iStar at AlphaFISH analytical resolution. Scale bar, 0.5 mm. (j–l) Quantification of spatial concordance between omics feature and HE feature. A tumor specification score is calculated by counting tumor-associated spots within a gridded split (0.03 mm^2^ square) of a biopsy tissue. Scatter plots show the Pearson correlation between H&E feature and that derived from **(j)** STrp (r=0.83), **(k)** SMib (r=0.81), and **(l)** combined STrp+SMib (r=0.84). **(m)** Cross-sample UMAP visualization of pooled STrp+SMib data from 102 biopsy sample pairs collected from 51 CRC patients. Tumor (pink) and nontumor (green) ROIs show well-separated distribution in UMAP space, suggesting distinct omics profiles. **(n)** Illustration of microbiological dysbiosis in CRC. (o–p) Quantification of microbiological diversity in tumor *vs*. nontumor biopsies. Shannon index **(o)** and Gini-Simpson index **(p)** are significantly lower in tumor samples. Analyzed by Kruskal-Wallis test, n=51. **(q)** Correlation of microbiological diversity metrics. A strong correlation (R2 = 0.97) is observed between the Shannon and Gini-Simpson indices across all samples. Tumor (pink) and non-tumor samples (green) enrich in different regimes, shaded bands represent 95% confidence and prediction intervals. For boxplots shown in panels h, o, and p, each box indicates the first quartile, median, and third quartile of the data; the whisker limit is 1.5 times the interquartile range; the black dot refers to a sample.

To confirm the nanostructured biochip’s capacity for the efficient extraction of bacterial RNAs, E. coli expressing EGFP was shown to release its RNA content after exposure to a nanoprobe array under >200g centrifugal force (**Suppl. Figure S3**). Scanning electron microscopy (SEM) confirmed successful physical interfacing between the nanoprobe array and biological samples, including both *Escherichia coli* bacteria and acute tissue sections (**Figure 2b**). Functional validation showed that only nanoprobes bearing 16S-priming sequences successfully extracted bacterial RNA, with RT-PCR amplification producing strong bands (400–500 bp) corresponding to the V3–V4 hypervariable region of the 16S gene (**Figure 2c**). Sensitivity testing using synthetic bacterial RNA fragments demonstrated detection at concentrations as low as 10⁻¹⁶ M (**Figure 2d**), with high reproducibility across biological replicates (n=5). Competitive binding assays using biochips functionalized with either poly(T), 16S-priming, or both baits (1:1 molar ratio, constant total molarity) confirmed the absence of cross-interference in dual-target capture, validated by statistical analysis (**Figure 2e**, two-tailed t-test, n = 9, ns).

The AlphaFISH platform was applied to 149 colorectal biopsy samples from 68 patients (**Figure 2f, g**), including paired tumor and adjacent non-tumor tissues from 51 CRC patients, and 47 samples from non-CRC controls. A biopsy tissue was trimmed into 5×5 mm^2^ small pieces and acutely interfaced with a biochip, which was specifically configured to profile 41 host transcripts, including mRNA involved in RNA methylation and inflammation responses (**Suppl. Table S1** and **Table S2**), and 51 microbial RNAs corresponding to 51 bacterial genera (**Suppl. Table S3**). Compared with random background gene sets, our 41 selected host transcripts demonstrate significantly higher activity scores and robust discriminative power in resolving diverse CRC-associated spatial phenotypes across all clinical subgroups (**Suppl. Figure S4**). This rapid touch-and-go interfacing with fresh tissue preserves the native mucosal microbiome. Quantitative analysis revealed that bacterial UMI counts were significantly enriched on the tissue surface compared to internal regions, whereas host mRNA UMI counts remained spatially consistent across both zones (**Suppl. Figure S5**). The quality of derived spatial omics data was assessed by quantifying the number of signal-positive ROIs, UMI counts per ROI for both host genes and bacterial genera, and the Shannon entropy of detected features (**Figure 2h**). These metrics confirmed consistent capture and high-complexity dual-omics data across large-scale biopsy samples and biochip formats (**Figure 2g, h**)

Functionally, the spatial maps of transcriptomic (STrp) and microbiome (SMib) signals aligned with H&E-stained histological features, and unsupervised clustering (UMAP) revealed distinct molecular domains (**Figure 2i**, more examples in **Suppl. Figure S6** and **Figure S7**). For individual biopsy samples, overlay of omics-derived tumor specificity with H&E-based annotations showed strong spatial correlation for STrp (r=0.83), SMib (r=0.81), and STrp plus SMib combined (r=0.84) analysis (**Figure 2j**-**l**). Across all CRC samples, a cross-sample UMAP analysis of 102 paired biopsies identified robust separation between tumor and non-tumor ROIs, underscoring AlphaFISH’s sensitivity to disease-associated omics patterns (**Figure 2m**). Importantly, AlphaFISH enables quantitative assessment of microbial dysbiosis using the derived spatial omics data. Tumor regions exhibited significantly reduced microbial diversity based on both Shannon and Gini-Simpson indices (p < 0.001, Kruskal–Wallis test, n = 51), with high concordance between indices (R² = 0.97). Tumors clustered consistently in the low-diversity regime, while non-tumor tissues retained higher ecological richness. These results demonstrate that AlphaFISH provides a robust, spatially resolved framework for dissecting host-microbiome interactions and uncovering disease-associated alterations in the tumor microenvironment.

### Computational architecture and performance of AlphaFISH

AlphaFISH is developed as a spatially aware transformer model to integrate dual-omics, spatial transcriptomics, and spatial microbiome data, enabling high-resolution tumor evaluation and host– microbiome interaction modelling in CRC. To achieve this goal, we analyzed 149 biopsies from 68 human subjects, comprising 51 CRC patients, 14 Crohn’s disease (CD) patients, and 2 healthy individuals (**Figure 3a**). These samples were distributed across both CRC and non-CRC cohorts. The biopsy samples were split into CRC tumor and nontumor samples, alongside CD and healthy controls (**Figure 3b, Suppl. Table S4** and **Table S5**), with pathological annotation in **Suppl. Tabel S6** and Tabel **S7**. Clinical TNM annotation revealed a range of tumor stages (**Figure 3c**).

**Figure 3.**
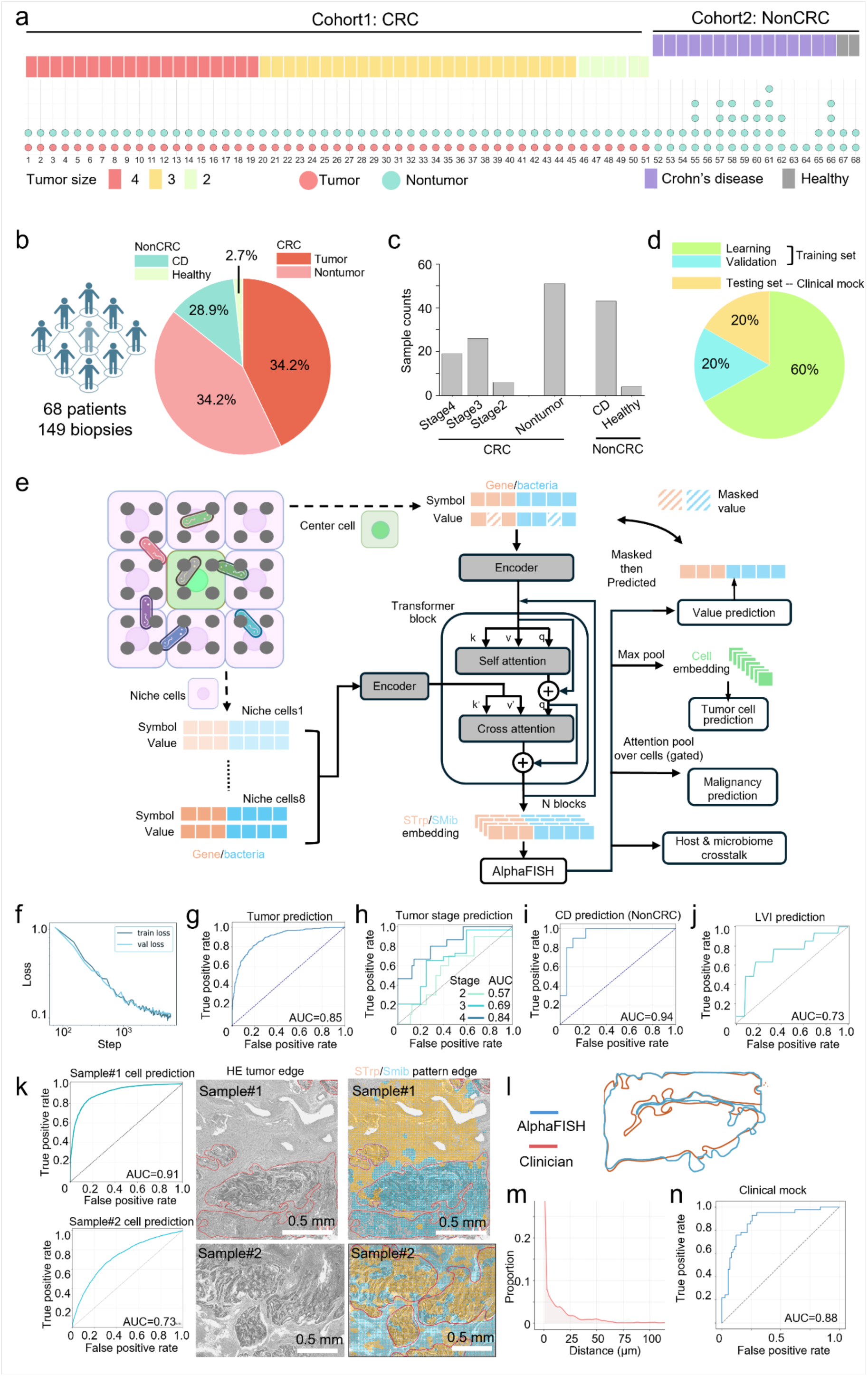
Computational architecture and performance of AlphaFISH. **(a)** Overview of the patient information. The study analyzes 149 biopsies from 68 human subjects in two cohorts: cohort 1 (CRC) includes 51 colorectal cancer patients, and cohort 2 (NonCRC) includes 14 Crohn’s disease (CD) patients and 2 healthy subjects. For each patient, colored squares indicate tumor stage, and colored circles indicate the preliminary clinical evaluation of the biopsy sample. **(b)** Pie chart showing the sample composition across 149 biopsies. **(c)** The counts of biopsy samples for different groups. **(d)** Data partitioning for machine learning and validation. The dataset is split into two sets: 80% for model training and 20% for model testing (unseen, clinical mock). The training set is further randomly split at a 3:1 ratio (learning vs. validation) during training. **(e)** Schematic of AlphaFISH architecture. Spatial tokenization: nanoprobes (gray dots) are aggregated into 4×4 bins to form quasi-single-cell units. For each unit, the local niche is defined as the center cell (green) surrounded by its eight immediate neighbors (purple, niche cell). The bacteria are scattered within the niche (rods). The AlphaFISH establishment is represented by STrp and SMib embeddings that effectively enable tumor cell prediction, malignancy scoring, and host–microbiome crosstalk inference via max pooling and attention pooling. **(f)** The training dynamics of AlphaFISH. Training loss (blue) and validation loss (yellow) decrease steadily and converge, confirming effective learning without overfitting. **(g–j)** Summary of validation performance by ROC curves, indicating the predictive accuracy on the validation set for **(g)** tumor malignancy (AUC=0.85), **(h)** tumor stage (discriminating Stage 4 with AUC=0.84), **(i)** CD differentiation from CRC (AUC=0.94), **(j)** LVI prediction (AUC=0.73). **(k)** Tumor cell prediction in two samples. Sample images showing tumor boundary evaluation by AlphaFISH at subcellular resolution using dual-omics spatial transcriptomics and spatial microbiome. Left: H&E image with clinician-annotated tumor edge (red line); right: AlphaFISH-predicted tumor cells (blue) *vs*. nontumor cells (yellow), overlaid HE image, showing strong alignment with histological tumor boundaries. Tumor boundary delineation by AlphaFISH or clinical annotation, scale bar 0.5mm. **(l)** Overlay of clinician-annotated (red) and AlphaFISH-predicted (blue) tumor boundaries. **(m)** Distribution of spatial distances between the predicted and clinician boundaries. **(n)** Diagnostic performance for the unseen clinical mock (AUC = 0.88).

To enable robust training and evaluation, the dataset was split into training and testing subsets, with 80% used for model training and 20% held out as a simulated clinical test set. Within the training set, a 3:1 ratio was applied to define learning and validation subsets (**Figure 3d**). AlphaFISH processes spatial data by aggregating nanoprobes into non-overlapping 4×4 bins, forming quasi-single-cell units. Each center cell is embedded along with its immediate neighborhood of eight surrounding cells, constructing a local niche. The model employs a dual-embedding strategy that encodes symbol–value pairs for both genes and microbes. A self-supervised masking scheme is applied to the center cell’s omics values, forcing the model to reconstruct missing information while maintaining full-fidelity inputs for the surrounding niche, thereby integrating both intrinsic and environmental signals.

At the core of AlphaFISH lies a stack of six transformer layers (**Figure 3e**). Detailed specifications of the model architecture, including embedding dimensions, number of attention heads, and specific training hyperparameters, are comprehensively listed in **the Supplement. Table S8**. The architecture first applies self-attention to learn intra-cellular dependencies within the center cell, followed by cross-attention that incorporates contextual embeddings from neighboring niche cells. This hierarchical encoding enables the model to evolve cell representations informed by both internal omics features and local spatial context. These refined embeddings are then used to infer multiple downstream tasks through pooling operations, max pooling, and attention pooling, yielding predictions for tumor cell identity, malignancy scores, and host–microbiome interactions. During training, both loss and validation metrics converged steadily, indicating effective learning and minimal overfitting (**Figure 3f**). The derived AlphaFISH model was found to contain distinct attention heads that address distinct pathological features, including tumor, tumor-adjacent, and nontumor. (**Suppl. Figure S8**). On the validation set, AlphaFISH achieved strong predictive performance across multiple tasks. Tumor malignancy scoring yielded an AUC of 0.85, while, achieving an AUC of 0.84 for stage 4 classification, AUC 0.73 for LVI and differentiated CD from CRC samples with an AUC of 0.94 (**Figure 3g-j**).

Additionally, AlphaFISH demonstrated high spatial fidelity in delineating tumor boundaries. When compared with expert pathologist annotations on H&E-stained sections, the model’s predicted tumor boundaries showed strong alignment (**Figure 3k-m**). Overlay analyses revealed close correspondence between predicted and annotated tumor edges, and spatial distance distributions confirmed concordance at subcellular resolution. These findings highlight AlphaFISH’s potential for automated histopathological assessment and its ability to resolve fine-grained tissue architecture. Notably, when tested on the held-out clinical mock set, AlphaFISH maintained high diagnostic accuracy, achieving an AUC of 0.88 in tumor cell classification (**Figure 3n**), underscoring its capacity for generalization to unseen patient data. Together, these results establish AlphaFISH as a powerful tool for spatially informed tumor detection and interpretation of host–microbiome interactions, capable of operating in clinically relevant settings with high accuracy and interpretability.

### Tumor-malignancy-associated spatial omics signature

To elucidate spatially resolved dual-omics signatures associated with CRC tumor malignancy, we used AlphaFISH to quantify the contributions of individual host genes and bacteria. Their importance scores were derived by ablating each feature from the input dual-omics vector (STrp + SMib) and measuring the drop in the malignancy prediction logits (Δlogits), with larger Δlogits indicating stronger influence (**Figure 4a**). Using this approach, AlphaFISH identified top-ranked microbial species with high effect sizes, revealing a subset of bacteria strongly associated with tumor malignancy (**Figure 4b**). Differential abundance analysis further confirmed that several of these bacteria, including *Fusobacterium*, were significantly enriched in tumor samples compared with paired nontumor samples (**Figure 4c**). Parallel analysis of host transcriptomics also reveals key genes with high impact on CRC malignancy status (**Figure 4d**). Genes such as *METTL3* showed both high model-derived effect size and significantly increased expression in tumor samples (**Figure 4e**). The spatial omics signatures were visualized using bubble plots, where bubble size reflects the percentage of quasi-single-cell units expressing a feature, and color indicates the average normalized abundance. The AlphaFISH-identified genes were projected onto a public single-cell CRC atlas (4.27 million cells from 650 patients), showing an enrichment in malignant epithelial and immune cell populations, including T and NK cells (**Figure 4f–g**).

**Figure 4.**
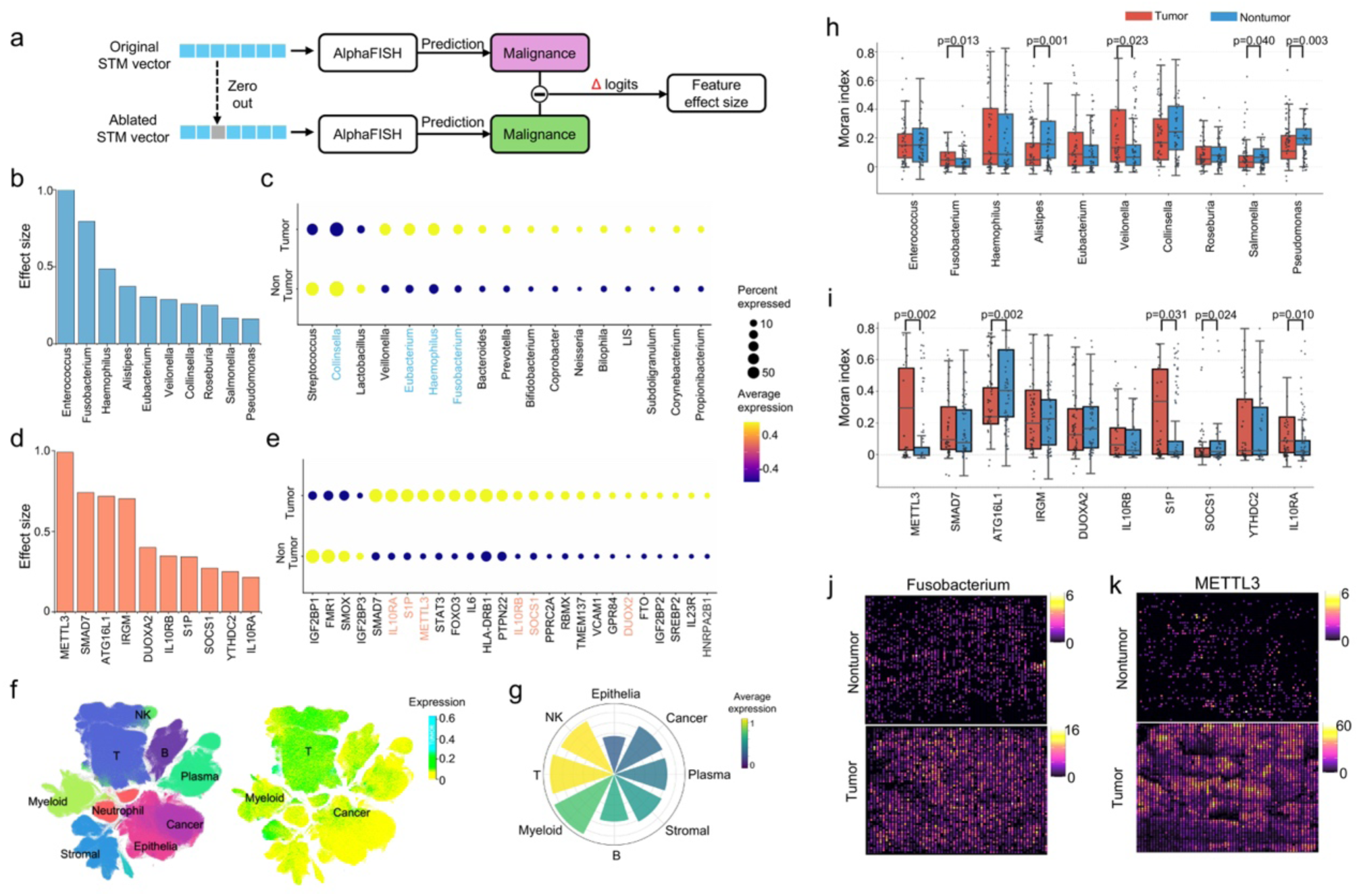
AlphaFISH identifies transcriptomic and microbial signatures associated with colorectal cancer malignancy. **(a)** The principle of scoring the importance of omics features in AlphaFISH. A malignancy score (logits) is derived from the STM (STrp+SMib) input vector. A specific feature within the vector is mathematically ablated; the effect size is quantified as a drop in predicted logits (Δlogits) between the original and ablated vector outputs: a larger drop implies a more critical role of the omics feature in association with tumor malignancy. **(b)** Microbial effect sizes of top bacteria ranked by AlphaFISH. **(c)** Differential bacteria abundance in tumor versus nontumor samples. **(d)** Transcriptomic effect sizes of top host genes ranked by AlphaFISH. **(e)** Differential gene expression in tumor versus nontumor samples. For **c** and **e**, the bubble plot shows the significant omics features (p<0.05) identified by SEURAT, analyzed by the Wilcoxon rank-sum test. Bubble size indicates the percentage of quasi-single-cell units expressing the gene or bacteria; color index represents the average of normalized abundance. Labels in blue **(c)** and orange **(e)** indicate the top-ranked bacteria or genes. **(f)** Projection of AlphaFISH-identified malignancy-associated genes onto a public single-cell CRC atlas comprising 4.27 million cells from 650 patients^27^. Left: UMAP space of major cell-type annotations for the public CRC atlas. Right: UMAP visualization of the normalized expression of the malignancy-associated genes identified by AlphaFISH. **(g)** Quantification of the enrichment of malignancy-associated genes across different cell types using the public atlas. In the polar bar plot, the radial distance indicates the ratio of cells expressing a specific gene set for a cell type; the color index indicates the average expression level of cells expressing the gene set. **(h–i)** Analysis of spatial autocorrelation for the omics features. Box plots displaying Moran’s index (MI) to quantify the spatial clustering or scattering of key bacteria **(h)** or host genes **(i)**. Comparison is made for Tumor (red) vs. Nontumor (blue) samples. P-values are calculated using the Kruskal-Wallis test. The black dots indicate an overlay of data points for individual samples; each box shows the first quartile, median, and third quartile. The whisker limit is 1.5 times the interquartile range. **(j–k)** Representative spatial expression (color indexed) of top-ranked omics features, Fusobacterium **(j)** and METTL3 **(k)**, in paired Tumor or Nontumor samples.

To assess the spatial context of the identified omics factors, we computed Moran’s index (MI) for the top bacterial and transcriptomic features. Tumor samples displayed significantly higher spatial autocorrelation for specific microbial (e.g., *Fusobacterium*) and host gene (e.g., *METTL3*) distributions compared to non-tumor counterparts (**Figure 4h, i**). Representative spatial maps further illustrated the clustering expression of these features in paired tumor versus nontumor samples, consistent with their elevated MI values (**Figure 4j, k**). Conversely, a reduction in spatial autocorrelation was observed for other features, such as *Alistipes* and *ATG16L1*, within the tumor microenvironment (**Suppl. Figures S9** & **S10**). These results demonstrate that AlphaFISH not only predicts malignancy with high fidelity but also identifies spatially structured microbial and transcriptomic signatures that are differentially enriched and clustered in CRC tumors, offering insights into tumor–microbiome–host transcriptomic interactions at subcellular resolution.

### CRC stage-specific spatial host-microbiome patterning

As illustrated in **Figure 5**, a comprehensive spatial multi-omics analysis of CRC reveals stage-specific host-microbiome interactions and associated immune remodelling across tumor progression. **Figure 5a** presents tumor size distributions in 51 CRC patients, stratified by tumor morphology (polypoid vs. infiltrative) and clinical stage (TNM stages 2–4). These measurements provide a clinical morphological context for molecular profiling. Differential analysis of microbial genera and host genes (Figure 5b, c) identifies stage-associated features, with key markers prioritized by the AlphaFISH model. These features show distinct expression patterns across tumor regions, suggesting coordinated changes in microbial composition and host transcription during disease progression. The ranking of associative features highlights specific bacteria and genes that contribute most to disease-stage classification (**Figure 5d**).

**Figure 5.**
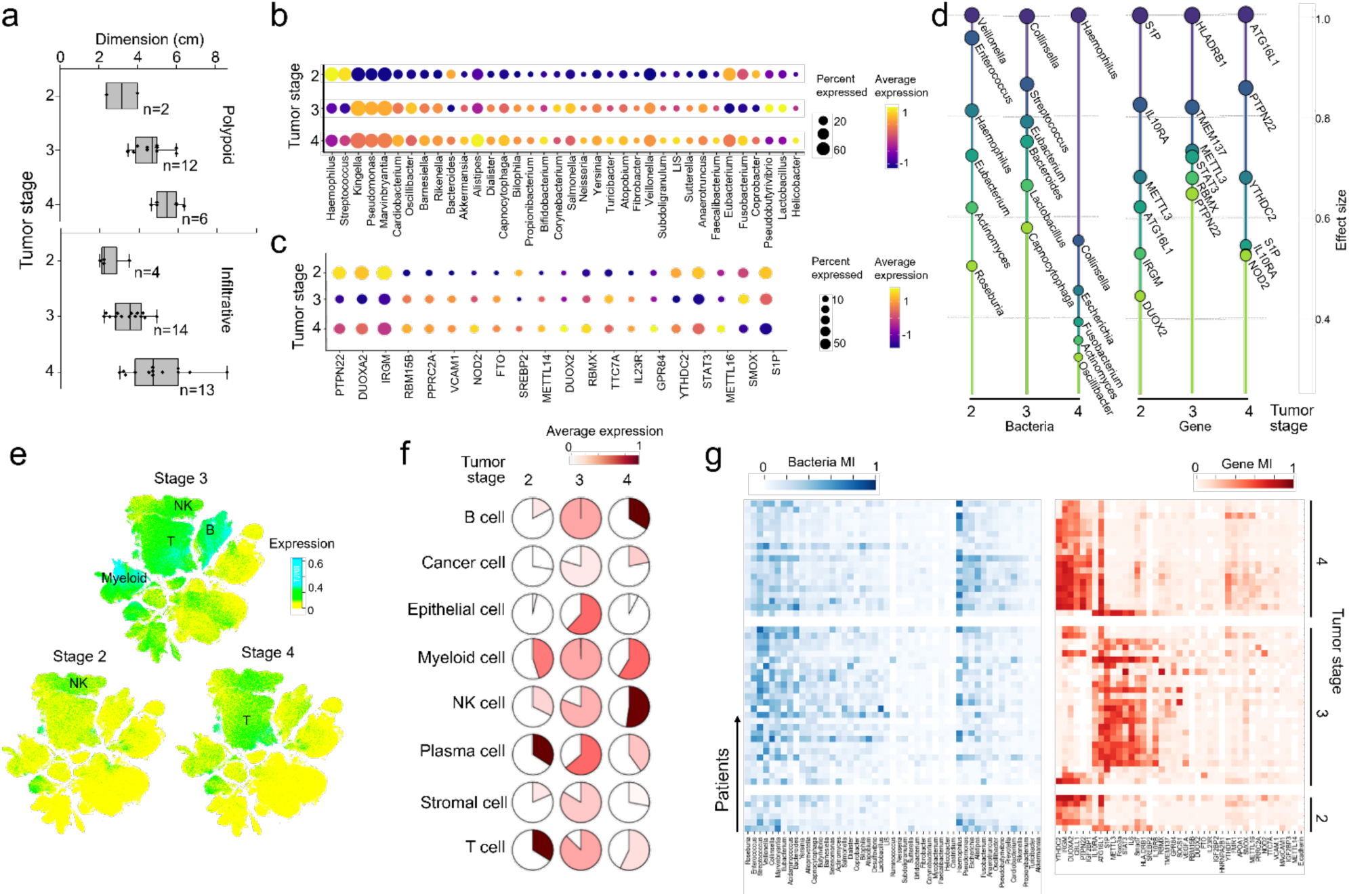
Stage-specific spatial microbiome-host signatures and immune landscape remodeling in colorectal cancer (CRC). (**a**) Box plots show quantification of tumor dimensions for 51 CRC patients with different tumor types (polypoid or infiltrative) across different TNM stages (2, 3, and 4). The number of patients for each condition is indicated. The black dots indicate an overlay of individual data points; each box shows the first quartile, median, and third quartile. The whisker limit is 1.5 times the interquartile range. (**b, c**) Analysis of significant differential abundance for genus-specific microbiome (**b**) and transcriptomic host genes (**c**) across different tumor stages. The calculation is performed using SEURAT; statistical analysis is performed using the Wilcoxon rank-sum test; only statistically significant bacteria genera and genes (adjusted p-value < 0.05) are visualized. Dot size represents the ratio of ROIs expressing the feature; color index represents average abundance. Orange-highlighted genus or blue-highlighted genes indicate those prioritized by AlphaFISH as key stage-associative features. (**d**) Top bacteria (left) and genes (right) ranked by AlphaFISH showing associative linkage to different tumor stages (2, 3, and 4) in CRC. Importance is quantified as an effect size, computed via feature ablation. For each feature (genus or gene), the model’s stage classification performance is measured with the feature present versus ablated (set to zero), and the difference in performance reflects its contribution. Effect sizes are then normalized across all features. (**e**) Projection of stage-associative genes (for stages 2, 3, and 4) onto the UMAP embedding of a public single-cell colorectal cancer cell atlas (4.27 million cells from 650 patients, hosted on CZ CELLxGENE). Each cell type is colored by the scaled aggregate expression score (green index) of the dominant gene list for the corresponding stage, computed as the normalized expression of the gene set for individual cells in the cell atlas space. Stage 2 associative genes are predominantly enriched in NK cells; stage 3 associative genes show broad enrichment across multiple immune cells (Myeloid, T, B, and NK cells); stage 4 associative genes indicate diminishing immune response from stage 3. (**f**) Quantitative characterization of stage-associative gene expression patterns in major cell types (panel e) using the public single-cell reference dataset. Pie charts show the ratio of cells in each cell-type space expressing at least one of the associative genes linked to the corresponding stage. Red index indicates the average expression abundance. Panels e and f show progressive remodelling of the immune microenvironment during tumor progression. (**g**) Joint analysis of spatial patterning for transcriptomics and microbiome across different tumor stages using Moran’s Index (MI). Heatmaps display the MI of all bacteria (left, blue) and genes (right, red) across 51 CRC patients of different stages (2–4). Higher MI indicates a clustered spatial distribution of relevant features.

To contextualize these transcriptomics-microbiome features within the immune landscape, we projected stage-associated gene sets onto a publicly available single-cell colorectal cancer cell atlas^27^ (4.27 million cells from 650 patients; **Figure 5e**). It was found that Stage 2-associated genes were predominantly enriched in natural killer (NK) cells, whereas Stage 3-associated genes mapped broadly across myeloid, T, B, and NK cell populations, indicating a coordinated immune activation. In contrast, Stage 4-associated genes exhibited reduced expression and cellular specificity, consistent with immune suppression in advanced CRC stage. Quantitative assessment further confirmed these trends, showing the proportion of cells expressing stage-specific gene sets and their average expression across major immune compartments (**Figure 5f**).

Different bacteria exhibit distinct spatial patterns associated with CRC tumor progression. For example, bacterial genera such as *Veillonella* and *Fusobacterium* exhibit a declining MI from stage 2 to stage 4, whereas *Escherichia* shows an increase at the late stage (**Suppl. Figure S11**). Host genes like S1P and IL10RA display a reduction, while IRGM and NOD2 show an increase in expression from stage 2 to stage 4 (**Suppl. Figure S12**). To assess the spatial evolution of transcriptomic and microbiome profiles, we computed Moran’s Index (MI) for all bacterial and host transcriptomic features across CRC patients at different tumor stages (**Figure 5g**). The joint Heatmap further reveals stage-dependent spatial patterning of the key host-microbiome features, showing elevated MI values indicating increased spatial clustering of selected features identified by AlphaFISH. These findings suggest that tumor progression is accompanied by coordinated spatial remodeling of both microbial communities and host gene expression, emphasizing the critical role of immune niches.

### AlphaFISH reveals immune–microbial crosstalk in colorectal cancer

Next, we used AlphaFISH-derived cross-attention maps to construct an interaction landscape between bacteria and host transcriptomic features. Normalized cross-attention weights were averaged across five independently seeded models to generate a robust interaction matrix, revealing distinct clusters of co-associated host genes and bacteria (**Figure 6a**). Leiden clustering of host genes identified eight transcriptional modules, each enriched for specific bacterial interactions. To explore the cellular context of these host gene modules, we projected the eight clusters onto the single-cell CRC atlas comprising 4.27 million cells from 650 patients^27^. UMAP analysis revealed distinct cell-type-specific expression patterns. For example, clusters 1 and 7 show enrichment in immune and epithelial/cancer cells, respectively (**Figure 6b**). Quantitative cell-type enrichment confirmed these associations (**Figure 6c**).

**Figure 6.**
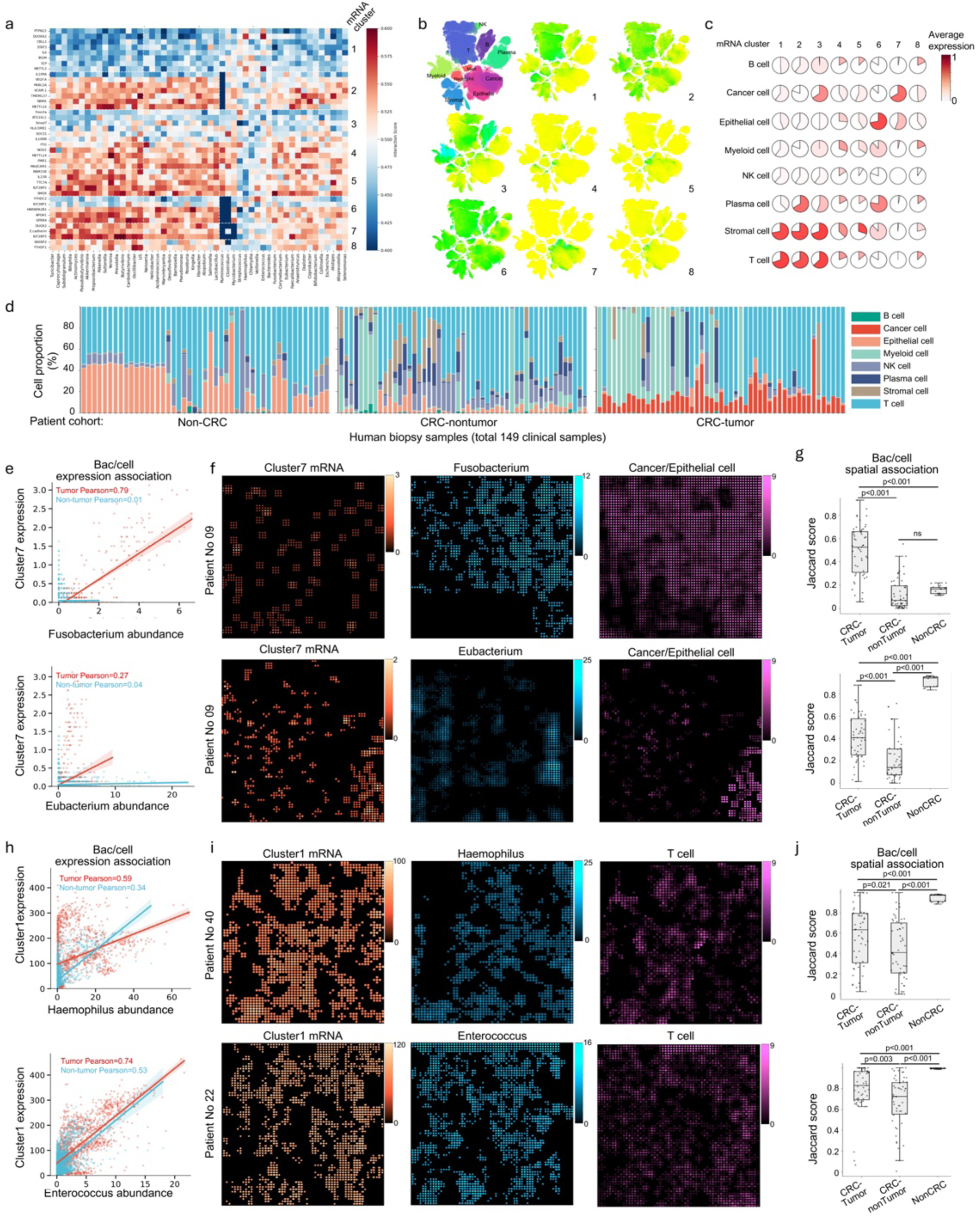
Key crosstalk factors of the immune-microbial niche in CRC. **(a)** Heatmap showing the host–microbiome interaction landscape in CRC. The indexed interaction scores between bacteria (columns) and host genes (rows) are derived from normalized raw cross-attention weights in AlphaFISH, averaged across five independently seeded models. Host genes are clustered into 8 distinct modules (clusters 1–8) via Leiden clustering. **(b)** Projection of the 8 gene clusters from (a) onto the UMAP of the public CRC single-cell atlas (4.27 million cells, 650 patients)^27^. Top-left corner: reference UMAP space of major cell type annotation for the public CRC atlas. Sub-panels 1–8 show UMAP visualizations of the normalized expression for the AlphaFISH-identified gene clusters. **(c)** Quantitative cell-type enrichment of the gene clusters (from **a** and **b**) using the public atlas. Pie charts show the ratio of cells in each cell-type space that express at least one gene in each cluster. The color index indicates the average expression abundance. **(d)** Nanoprobe-derived cell-type composition across different biopsy samples of different patient cohorts (149 biopsies). Stacked bar chart showing the proportion of different cell types in the biopsy samples for NonCRC (n=47), CRC-nontumor (n=51), and CRC-tumor (n=51). Cell type classification is determined from AlphaFISH-derived quasi-single-cell profiles using a classifier trained on the public CRC atlas. (**e–g**) Spatial association between *Fusobacterium*/*Eubacterium* and the cancer/epithelial cell-associated omics features. **(e)** Scatter plots showing the co-expression correlation between the *Fusobacterium* (top) or *Eubacterium* (bottom) and cluster 7 genes (cancer/epithelial-associated) within the localized CRC niche. Both *Fusobacterium* and *Eubacterium* show positive colocalized correlation only in tumor biopsies, with *Fusobacterium* exhibiting *the highest* Pearson correlation (r=0.79). **(f)** Representative spatial mapping of: cluster 7 expression (orange), bacteria abundance (cyan), and cancer/epithelial cells (magenta) on a biochip. Color index indicates the mean value expression or abundance. **(g)** Quantification of spatial co-localization between the expression patterns of bacteria and cancer/epithelial cells, indicated by the Jaccard index. **(h–j)** Spatial association between *Haemophilus*/*Enterococcus* and immune cell-associated omics features. **(h)** Scatter plots showing the co-expression correlation between the *Haemophilus* (top) or *Enterococcus* (bottom) and cluster 1 genes (immune cell-associated) within the localized CRC niche. *Haemophilus* (r=0.59) and *Enterococcus* (r=0.74) show highly positive, colocalized correlations, without differentiation between tumor and nontumor status, indicating their association with immune activation. **(i)** Representative spatial mapping of: cluster 1 expression (orange), bacteria abundance (cyan), and T cells (magenta) on a biochip. **(j)** Quantification of spatial co-localization between the expression patterns of bacteria and T cells, indicated by the Jaccard index. For **g** and **j**, the Jaccard Index is calculated as the ratio of intersection spots to union spots for bacterial and cell omics patterns—P-values determined by Mann-Whitney U-test. The black dots indicate an overlay of individual data points; each box shows the first quartile, median, and third quartile. The whisker limit is 1.5 times the interquartile range.

Using nanoprobe-classified quasi-single-cell profiles, we analyzed cell-type composition across the 149 biopsy samples from three patient cohorts (NonCRC, n=47; CRC–nontumor, n=51; and CRC– tumor, n=51), and found that CRC tumors show increased proportions of cancer-like cells, while immune cell representation remains a key variable (**Figure 6d**). We then assessed spatial co-localization between bacteria and host gene modules. *Fusobacterium* and *Eubacterium* exhibited strong spatial correlation with epithelial-associated cluster 7 genes exclusively in tumor biopsies, with *Fusobacterium* showing the highest correlation (Pearson *r* = 0.79) (**Figure 6e**). Spatial maps visualized the tri-layered co-localization of bacteria, host gene expression, and cancer cells (**Figure 6f**), further supported by significantly elevated Jaccard indices in tumors compared to non-tumors (**Figure 6g**). In contrast, *Haemophilus* and *Enterococcus* showed high spatial correlation with immune-associated cluster 1 genes across both tumor and nontumor samples (**Figure 6h-j**), suggesting a more general association with immune activation and revealing distinct spatially resolved host–microbe interactions in CRC niches beyond the well-documented *Fusobacterium* association with tumor epithelium^3,28^.

### Prediction of post-resection tumor recurrence

To translate our spatial omics and tumor microenvironment observations into a clinically applicable prognostic framework, we conducted a prospective analysis on a longitudinal cohort of CRC patients with clinical follow-up covering one-year period post tumor resection (**Figure 7a**). We aligned foundational clinical pathological features across the cohort to chart a comprehensive landscape of standard risk markers (**Figure 7b, Suppl. Figure S14**). Long-term disease progression was defined as a composite outcome of tumour recurrence or metastasis. When AlphaFISH was tasked to predict this outcome, it achieved an area under the curve (AUC) of 0.81. This performance exceeded that of a baseline consensus framework derived from conventional clinical matrice acquired at the surgical operation stage, which comprised lymphovascular invasion (LVI), perineural invasion (PI), tumor budding and the Ki-67 index and yielded an AUC of only 0.45 (**Figure 7c, Suppl. Table S6, S7** and **Figure S15**). This supurior performance suggests that the dual-omics spatial architecture harbors complex, deeply embedded prognostic signals that extend far-beyond standard histopathological staging.

**Figure 7.**
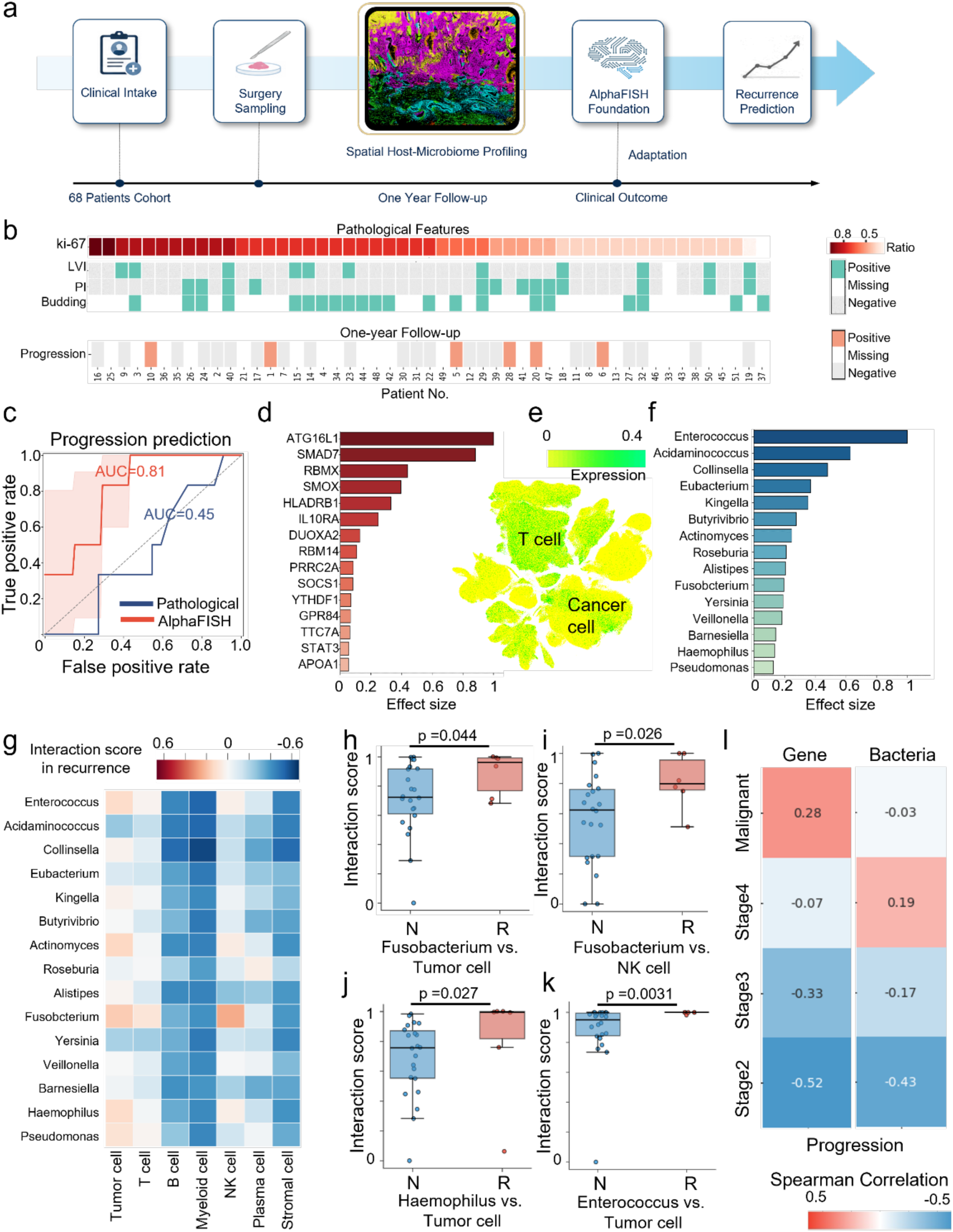
AlphaFISH predicts post-resection tumor recurrence in colorectal cancer (CRC). **(a)** Study design and clinical timeline for the CRC patient cohort, tracking disease progression related to tumor recurrence or metastasis over a longitudinal one-year follow-up after surgical tumor removal. **(b)** Pathological landscape of the cohort (n = 51). The heatmap aligns foundational pathological features, including Ki-67 expression, lymphovascular invasion (LVI), perineural invasion (PI), and tumor budding, with the actual occurrence of disease progression. **(c)** Receiver operating characteristic (ROC) analysis evaluating the predictive performance for long-term disease progression. The AlphaFISH model (red; AUC = 0.81), the traditional clinical consensus model (blue, integrating LVI, PI, Ki-67, and tumor budding; AUC = 0.45). **(d)** Feature ablation analysis ranking the top dominant host genetic features driving the model’s predictive accuracy for disease progression. **(e)** Anchor of the top-rank prognostic host genes to the scRNA-seq atlas covering 4.27 million cells across 650 patients^27^, demonstrating preferential enrichment of T cells and malignant epithelial populations. **(f)** Top dominant microbiome contribution for recurrence prediction. **(g)** Heatmap profiling the subtraction score of niche-level spatial interactions between clinical outcomes (Δ*S* = *S_Re_* − *S_NonRe_*) across the top 15 bacterial genera (rows) and 7 major cell types (columns). Red indicates enriched spatial crosstalk in progressing tumors, while blue denotes depletion. **(h-k)** Micro-niche interaction scores are stratified by clinical outcome (N: Normal follow-up; R: Recurrence, n = 28). Signatures are shown for (h) Fusobacterium vs. Tumor cell (i) Fusobacterium vs. NK cell, (j) Haemophilus vs. Tumor cell, and (k) Enterococcus vs. Tumor cell. The p-values are determined via Wilcoxon sum rank test. Box limits indicate the 25th and 75th percentiles, center lines denote the median, and whiskers extend to 1.5× interquartile range (IQR), with individual patient data points superimposed. **(l)** Spearman correlation matrix tracking the clinical correlation of dominant host gene expressions and bacterial abundances across progressive pathological states.

To decipher the pathological architecture driving AlphaFISH’s predictive performance, we examined the model’s internal feature importance and conducted feature ablation analyses to isolate dominant host transcriptomic and microbial determinants. Within the predictive framework, feature importance ranking identified a core set of host genes, including *ATG16L1*, *SMAD7*, *RBMX*, and *SMOX*, as the leading contributors to the model’s risk output (**Figure 7d**). These top-rank host genes were anchored and projected onto the reference single-cell RNA-sequencing (scRNA-seq) datasets covering 4.27 million cells across 650 patients^27^, revealing that these transcriptomic signatures were predominantly converged to T cells and malignant epithelial cancer cells (**Figure 7e**). Meanwhile, modeling of the intra-tumor microbiome by AlphaFISH pinpointed *Enterococcus*, *Eubacterium*, *Fusobacterium*, and *Haemophilus* as dominant microbial factors for patient stratification (**Figure 7f**). These genera have previously been closely linked to specific malignant micro-niches and tumor stages^29–31^. To study the precise multicellular axes driving AlphaFISH’s predictions, we differentiated the spatial interaction landscape between progressing and non-progressing patients. By evaluating the subtraction scores of niche-level spatial interactions (ΔScore), we found that tumors from progressing patients harbor an ecosystem-wide enrichment of highly specific bacterial-cellular crosstalks (**Figure 7g**). Aggregating these niche dynamics into patient-level validations, we identified multiple spatial interactions that were robustly elevated in the recurrence group (**Figure 7h-k**), including significant spatial co-localization of *Fusobacterium* and Tumor cells (p = 0.044), *Fusobacterium* and NK cells (p = 0.026), *Haemophilus* and Tumor cells (p = 0.027;), and *Enterococcus* and Tumor cells (p = 0.0031). Longitudinal stage alignment confirmed that these dominant host-microbiome topological features were highly correlated with progressive pathological states from early to advanced malignancy (**Figure 7l**). Taken together, these data demonstrate that the dense spatial co-localization of specific pathogenic microbiomes and tumor/immune cells within compartmental micro-niches represents a potent, clinically predictive multi-omics axis that decodes prospective patient prognosis.

## Discussion

In this study, we present AlphaFISH, a platform technology integrating both technical and computational innovations for multi-omics spatial analysis of acute clinical biopsies at subcellular resolution (4 um). The system uses a sequencing-free, high-throughput, dual-omics spatial profiling technique to construct, to date, the largest spatial transcriptomics and spatial microbiome datasets acquired from human subjects. Deep learning of the data enables the development of a transformer model with joint embeddings of gene expression, spatial architecture, and the microbial microenvironment in colon tissues, achieving over 90% accuracy in predicting CRC-associated pathological features using unseen spatial omics inputs. AlphaFISH also deciphers novel spatial phenotypes underlying CRC progression, revealing natural killer cell activation as a marker of tumor malignancy and uncovering the spatial patterning of key bacterial and immune crosstalks. The results particularly emphasize the interactive association between *Fusobacterium* and cancer cells, as well as the underappreciated linkage between *Haemophilus* and immune cells.

The field of multi-omics spatial profiling has seen rapid advances, driven by innovations in spatially resolved sequencing, high-resolution imaging, spatial barcoding, and integrated data analysis^19,32^. These advances hold great promise for advancing precision disease management by data-driven, tissue-aware decision-making that enhances diagnostic accuracy, guides treatment selection, and supports the discovery of novel spatial biomarkers across multiple disease contexts^33–35^. A pioneering study by Jia et al. presents an AI-driven spatial transcriptomics system, which integrates spatial patterns of immune-related gene expression to predict hepatocellular carcinoma recurrence by leveraging machine learning on spatial omics data^36^. This work underscores the clinical potential of AI-powered interpretation of spatial multi-omics by deciphering the inherent complexity, high dimensionality, and spatial structure of the data. However, the prospect of integrating spatial omics data with AI faces numerous challenges, one of which is the lack of large-scale, high-quality spatial omics datasets for the development of robust AI models^13,37^. Spatial omics remains relatively data-poor: most existing datasets are limited in sample size, tissue types, and patient diversity, restricting the AI model’s ability to learn complex biological patterns that are representative across populations and disease contexts^13,19^. Moreover, spatial omics data are often generated using different platforms^15,22^, each with varying resolutions, gene panels, and noise profiles, further complicating data harmonization and model transferability. For AI models, especially deep learning architectures that require large training sets, this lack of scale leads to overfitting, poor reproducibility, and limited clinical relevance. Moreover, the scarcity of well-annotated labels (e.g., cell types, disease states, clinical outcomes) in association with existing spatial omics data hampers the development of supervised models.

To unlock the full potential of AI in spatial omics clinical applications, this study constructs the largest clinical spatial multi-omics data library to date, marking a significant milestone in translational spatial omics and AI-driven discovery. By profiling 149 colorectal biopsies from 68 human subjects of CRC-, CD-, and healthy cohorts, this scale is aimed to capture a uniquely comprehensive view of spatial tissue organization across inflammatory, cancerogenic, and normal states, that includes of both immune and microbial features alongside host transcriptomics, providing an unprecedented opportunity to decode the complex cellular and molecular interactions within the colorectal microenvironment. Importantly, the scale and diversity of this dataset address a major limitation that has hindered the development of robust machine learning models in spatial omics by integrating spatial resolution with pathological and disease-specific variation. This resource enables the training of AlphaFISH models that generalize across patient populations and tissue contexts and lays the groundwork for discovering the spatial signatures associated with host–microbiome crosstalk in CRC. Towards the clinical application of spatial omics, this data library is not only a technical achievement but also a foundational demonstration of how to build reliable and interpretable AI tools for spatial omics-based precision medicine.

As the technical core, the establishment of AlphaFISH relies on the co-detection of eukaryotic and prokaryotic RNAs within the same spatial assay, which has been a significant challenge due to fundamental differences in RNA biology and current technological limitations^11^. Most spatial transcriptomics platforms rely on poly(dT) capture, which efficiently targets polyadenylated eukaryotic mRNAs but fails to capture prokaryotic RNAs, which generally lack poly(A) tails^38^. Microbial RNA is often low in abundance, fragmented, and dominated by ribosomal RNA, making it difficult to detect informative microbial transcripts^38^. Designing probe panels that can simultaneously and specifically target diverse bacterial species and human transcripts is complex, given the high sequence variability and limited annotation of many microbial genomes^11^. Spatial resolution and sensitivity are also limiting factors—microbes may be sparsely distributed within tissues, and overlapping signals from nearby host and microbial cells can be difficult to resolve accurately due to differences in expression dynamics and transcript structure^7^. Furthermore, tissue preservation methods, such as formalin fixation, can degrade microbial RNA or alter the microbiota, complicating spatial localization. These challenges highlight the value of the technical innovation of AlphaFISH, featuring specialized protocols, dual-target probe systems, and robust bioinformatic frameworks to enable accurate cross-organism spatial omics profiling of transcriptomics and microbiome in clinical biopsies.

Built on these technical and computational innovations, our study provides compelling evidence that spatial host–microbiome interactions are intricately linked to immune niche remodelling in CRC. Using AlphaFISH, we identified distinct clusters of co-associated host genes and microbial taxa, revealing structured crosstalk across microbial and immune components. It was not surprising to find a spatial correlation showing that *Fusobacterium* and *Eubacterium* are exclusively co-localized with epithelial/cancer-associated omics features in CRC tumor biopsies, highlighting their well-documented roles in shaping malignant niches. In contrast, *Haemophilus* and *Enterococcus* showed strong associations with immune-related gene modules (cluster 1) across both tumor and nontumor samples, suggesting a broader, coordinated role in immune activation. Moreover, cell-type deconvolution reveals increased cancer-like epithelial cell composition and variable immune cell distributions across CRC samples and tumor stages, further supporting a dynamic tumor–immune– microbe interface. This work underscores the importance of spatial context in understanding host– microbe crosstalk, revealing not only the cancer-specific enrichment of certain bacterial genera but also their coordinated interaction with distinct immune cell states. These findings position AI-facilitated spatial multi-omics as a powerful framework for dissecting the ecological complexity of the tumor microenvironment and identifying immune-modulating microbial targets in CRC.

Meanwhile, we acknowledge several limitations in this study. First, increasing the throughput of transcriptomic and microbiome profiling would enhance the analytical depth of spatial omics studies. In the current AlphaFISH platform, we simultaneously profiled 41 host transcripts and 51 bacterial genera, which is relatively small compared to sequencing-based approaches. While this dual-omics design provides foundational insights into host–microbe interactions, increasing the number of detectable targets would enable broader and more nuanced pathological discovery. A higher number of host transcripts could capture rare cell states and regulatory pathways, while an expanded microbial panel would improve detection of low-abundance or disease-specific bacterial taxa. Within a similar technical framework, target coverage at a few hundred would be achievable and further strengthen the resolution and interpretability of spatial ecological dynamics within complex tissue environments, such as colorectal tumors. Second, current data labelling primarily relies on TNM stage information, a clinical measure of tumor progression. To enhance the utility of spatial omics for precision medicine, future expansions should incorporate additional treatment-related metrics, such as recurrence, response to immunotherapy, and survival. Linking spatial omics features with these endpoints would allow the identification of predictive biomarkers and spatial phenotypes associated with therapeutic efficacy or resistance. Such integration would significantly improve disease stratification and guide personalized treatment decisions, transforming spatial omics from a descriptive tool into a clinically actionable framework for patient management. Thirdly, the deep learning architecture remains intrinsically constrained by the informational content of the pre-selected molecular anchors. Advances in generative AI and large-scale biological foundation models promise inferring of whole-transcriptome landscapes from sparse molecular inputs^39^. These emerging models leverage vast pre-trained knowledge to perform high-precision imputation based on limited data points^40^. By treating our targeted spatial anchors as seed features, future iterations of AlphaFISH could potentially reconstruct genome-wide expression patterns *in silico*, enabling both cost-efficiency and greater information richness through generative inference.

In conclusion, our study presents AlphaFISH to demonstrate the power of AI-integrated, high-resolution spatial multi-omics to overcome clinical data limitations and to enable accurate CRC pathology prediction, revealing dynamic tumor–microbe–immune interactions through disease progression. This platform sets a benchmark for spatial omics-based prognostics and provides a scalable foundation for precision medicine in complex tissue ecosystems.

## Methods

### Clinical study

All procedures involving human samples were performed in accordance with the ethical standards of the institutional and national research committees, as outlined in the 1964 Helsinki Declaration and its later amendments, or comparable ethical standards. Ethical approval (2024ZSLYEC-304) with informed consent was obtained at the Sixth Affiliated Hospital of Sun Yat-sen University.

68 Human subjects were recruited at the Sixth Affiliated Hospital of Sun Yat-sen University and included 51 CRC patients, 15 CD patients, and 2 healthy subjects. The researchers have received the necessary training for the research program and have a sufficient understanding of the research. Dedicated quality control personnel check the clinical execution to ensure that all the trial conditions meet the research and clinical requirements of the program. Throughout the study, the researchers conduct the clinical operation according to the standard principles and procedures outlined in the approved research protocol. The corresponding project documents were achieved and preserved by the quality control personnel. The quality assurance department of the clinical research unit (Sixth Affiliated Hospital of Sun Yat-sen University) audits the clinical research.

The nature, objectives, procedures, expectations, timing, potential risks and benefits, and any discomfort that may arise from the operation have been explained in detail to each subject with informed consent. Each subject was informed that their participation in the study was voluntary, that they could withdraw at any time, and that withdrawal would not affect their subsequent treatment.

### Fabrication and functionalization of nanostructured biochips

Biochips with silicon nanoprobes were prepared using a top-down MEMS approach. An array of dots (0.8 μm in diameter, spaced by 4 μm center-to-center) was produced by photolithography on a silicon wafer. Next, deep reactive ion etching (DRIE) was performed to fabricate an array of micropillars (∼10 μm height), which were further processed by thermal wet oxidation and chemical etching to reduce the diameter to around 0.8 μm. The silicon wafer with the nanoprobes was then cut into square chips (1×1 mm2) using a UV laser (**Suppl. Figure S1**).

For functionalization, the nanoprobe biochips were processed with piranha solution (v/v = 3:1, 98% H_2_SO_4_:27.5% H_2_O_2_) at 90 ℃ for 90 min to produce surface hydroxy, and subsequently rinsed with deionized water, methanol, methanol/dichloride methane (DCM) mixture (v/v = 3:1). The chips were immersed in DCM solution containing 3% (v/v) (3-aminopropyl)triethoxysilane (APTES) for 3 hours to render amine groups on the nanoprobe surface. Afterwards, the chips were rinsed with ethanol, isopropyl alcohol, and deionized water. The amine groups were then activated with glutaraldehyde (15%, v/v) for 2 hours, followed by crosslinking with the functional protein streptavidin (SA, for mRNA chip, 10 μg/ml) for 2 hours. The chips were then placed in 1% BSA solution (in PBS) containing 0.1% Triton X-100 for 5 hours to block unreacted groups. The biochips were further reacted with biotinylated polyT sequences (100 nM) and biotinylated 16S bait sequences (100 nM) for 1 hour, followed by incubation of biotin (0.1 mg/ml) for another 2 hours to block the unreacted binding sites on the SA proteins. All reactions were performed at room temperature unless otherwise specified.

### Multiplexed encoding and decoding for on-chip transcriptomic analysis

For dual-omics sampling, fresh-frozen biopsies underwent two freeze-thaw cycles before being profiled with the nanoprobe biochip. The tissue-biochip interface was further spun at 300 × g for improved contact. The biochip was then removed from the tissue for subsequent processing. For multiplexed encoding and decoding of AlphaFISH-extracted RNA transcripts (mRNA of host and 16S rRNA of bacteria), on-chip hybridization with targeted RNA transcripts was performed by adding DNA-encoded reporter sequences, which were also fluorescently labeled with specific Spectrum-CODE^@^ beads for visualization in later confocal microscopy. The Spectrum-CODE beads were commercially acquired (300-400 nm in diameter, ANRim Biotech). A 7-channel Spectrum-CODE^@^ system was used. Each Spectrum-CODE bead was pre-conjugated with a unique fluorophore combination selected from the following 7 fluorophores: quantum dot 547 nm (Qdot547), Qdot572, Qdot604, Qdot641, AlexaFluor488, AlexaFluor552, or AlexaFluor638, providing a digitized spectrum that can be used to decode the RNA transcript by fluorescence imaging. In this framework, “1” denoted inclusion and “0” exclusion of a fluorophore on a Spectrum-CODE bead, providing a 7-digit spectral code for a single RNA transcript. In this way, the 7-channel fluorescent system is expanded to encode more than 100 transcripts (maximum capacity: 27) in a single imaging cycle (**Suppl. Figure S2**). The Spectrum-CODE beads were dispersed in a 1% BSA solution containing 0.1% Triton X-100 and stored at 4 ℃ for further use.

### Fluorescence microscopy

The fluorescence images were captured with a confocal laser-scanning microscope (Leica SP8, 63× oil-immersion objective, NA 1.4, Leica Microsystems). Briefly, after omics sampling, the biochips were removed from the contacting tissue and placed in RNALater (Beyotime). Before imaging-based analysis, the DNA-encoded reporter sequences were incubated with the biochips for 30 minutes, followed by an extensive PBS rinse. Fluorescence images were acquired using a confocal laser scanning microscope. All imaging parameters, including laser power, gain, and scanning speed, were fixed throughout the microscopy process. Scanning was performed with a 1 μm step size along the Z-axis spanning the whole layer. For data collection from a biochip, imaging-based decoding was performed using spectrum encoding (**Suppl. Table S1** and **S3**). To obtain a complete view of a biochip, we used the navigator mode in Leica Microsystems to capture over 200 tiles. The resulting large-scale tiles were merged using Leica Application Suite X (LAS X).

### Generation of STrp and SMib

To transform the fluorescence image into a matrix, ImageJ, Python, and R were used. The identification of Spectrum-CODE beads was performed in Python. For analyzing the expression of an RNA transcript, the fluorescence image of a biochip undergoes processes including binarization, mask generation, nanoprobe ROI selection, denoising, and spectral vector composition, by extracting signals from all 7 fluorescence channels. The featured spectral vectors were decoded using a machine learning model to derive the Spectrum-CODE associated with the nanoprobes, which were then counted to indicate the molecular abundance of different transcripts. The machine learning model was pre-trained on a dataset of actual fluorescence patterns collected across all possible Spectrum-CODEs. For each, >200 samples were collected across multiple biochips to construct the training dataset.

Specifically, the fluorescence images were acquired as a 3-dimensional stack (*x*, *y*, *z*), which were cropped in ImageJ to remove unnecessary edges, and then loaded in Python using the OpenCV package. For each nanoprobe ROI, the associated Spectrum-CODE puncta were identified using the connected_components function in Python; these were then recognized by randomforest to determine the corresponding Spectrum-CODE, which was then counted. Counts for all 91 Spectrum-CODEs associated with each nanoprobe across the entire biochip were used to compose the RNA expression matrix for the AlphaFISH tissue sampling site, which was later subjected to spatial transcriptomic analysis.

### Histological feature extraction via iStar

Histological features were extracted from H&E-stained images using the iStar framework^41^. To align with the AlphaFISH spatial resolution, H&E images were virtually sampled using a sliding window approach with a stride of 4 μm. At each coordinate, circular regions of interest (ROIs) with a diameter of 0.8 μm were defined to match the AlphaFISH spot geometry. The H&E images were rescaled to standardize pixel size and partitioned into hierarchical image tiles. Subsequently, a Vision Transformer (ViT) was employed to extract high-dimensional morphological features from each sampling spot, generating a histology embedding map spatially registered to the multi-omics data.

### Model architecture of AlphaFISH

#### Data preprocessing

To define spatial units for model training, raw spot-level gene counts and microbe counts were aggregated within each sample. Here, spots denote the original sampling locations, gene counts denote host gene-expression measurements, and microbe counts denote genus-level microbial measurements. Counts were summed across non-overlapping 4×4 spot bins, which were used as the model units because their physical scale better approximates cellular-scale tissue neighborhoods than individual spots. We use bin below to refer to these 4×4 aggregated units. Bins with any missing spot were excluded. For each retained bin, the local spatial information was defined as the center bin and its eight nearest neighboring bins on the spatial grid. A bin was used as a training center only when all eight neighboring bins were present and the center bin contained at least five non-zero gene or microbe features. Counts were transformed as log(1 + x) to reduce the influence of highly abundant genes and microbes.

#### Feature encoder

AlphaFISH used a feature encoder followed by a transformer backbone. For each bin, the encoder mapped gene identities, microbe identities and their abundance values into token embeddings. The transformer updated the representation of the center bin by attending to features from that bin and from its eight neighboring bins. To provide the transformer with one token per observed feature, each non-zero gene or microbe in a bin was converted into a vector representation. For a bin with feature identity and value pairs {(*g_i_*, *v_i_*) or *i* = 1, . . . , *L* , where *v_i_* is the log1p-scaled abundance, the input embedding for feature *i* was defined as

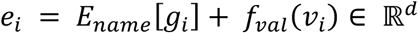

Here, *E_name_* is a learnable embedding table for feature identity. The abundance values in AlphaFISH input are continuous non-negative scalars. We therefore used the continuous value embedding *f_val_*(*v*) rather than assigning abundance values to discrete bins.

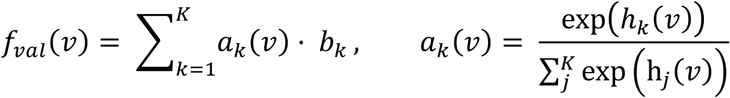

The basis embeddings *b_k_* defined a learnable value basis, and the coefficients *a_k_*(*v*) were softmax-normalized scalar functions of the abundance value. This design was motivated by previous works{Hao, 2024 #124;Cao, 2024 #125}, which reported improved performance for continuous expression-value encoding compared with discretization-based schemes.

#### Transformer block

The Transformer block served as a core component of AlphaFISH. It could capture both intra-cell and inter-cell interactions. Let *X* ∈ ℝ*^L^*^×*d*^ be the gene/microbe feature matrix of the center quasi-single-cell (generated by the above feature encoder) and *Y* ∈ ℝ*^L^*^′×*d*^ the concatenated features from its eight neighbors. *L* and *L*^0^ are the lengths of non-zero features in a center cell and its eight niche cells. *d* is the embedding size. We used two types of attention, self-attention and cross-attention, to separately capture intra-cell and inter-cell interactions. Self-attention was performed in the following way:

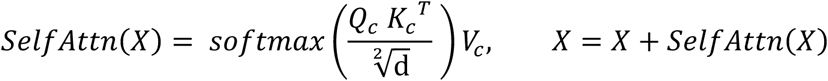

Here, the center cell feature *X* is converted to *Q_c_*, *K_c_* and *V_c_* by three different learnable functions, and is updated in a residual manner. Cross-attention was performed in the following way:

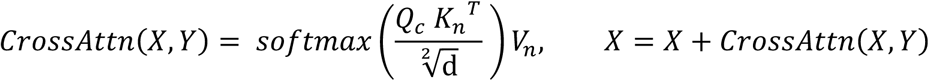

Here, *Q_c_* is generated from the center cell feature *X*, the others, *K_n_* and *V_n_* are generated from niche cell features *Y*. In this way, center cells and their niche cells can communicate with each other. We used *N* = 6 transformer blocks to perform repeated information exchange between the center cells and niche cells.

#### Pretraining: masked value reconstruction

To learn spatial gene and microbe representations without downstream task labels, AlphaFISH was pretrained with masked value reconstruction. In each center bin, 20% of feature values were masked. The values of the eight neighboring bins were not masked and were provided as complete spatial context. The prediction head regressed the original log1p-scaled abundance values at the masked center-bin positions using mean squared error.

#### Training protocol

The backbone was optimized with AdamW (learning rate, 1 × 10^−4^). The schedule used a linear warm-up over the first 10% of training steps, followed by learning-rate decay. Dropout was set to 0.1, and gradients were clipped by global norm at 0.5. Training used fp16 mixed precision, a batch size of 256 and 50 epochs on one NVIDIA A100 GPU with 80 GB memory. Random seeds were fixed for data partitioning and data-loader shuffling.

### Downstream tasks

#### Tumor cell identification

To train local tumor-annotation models, expert H&E annotations were transferred to matched spatial transcriptomics spots in the HE-ST samples. The two HE-ST groups were sampled from different patients. H&E images were used only for annotation transfer and tissue visualization, not as input channels to the AlphaFISH transformer. Bins whose 4×4 spot regions intersected the pathologist-annotated lesion boundary were excluded from supervised training and evaluation to reduce boundary-label ambiguity. For bin-level tumor classification, embeddings from the frozen pretrained backbone were used as node features in a weighted k-nearest-neighbor tissue graph. Each bin was connected to its 25 nearest bins by Euclidean distance in spatial coordinates, and neighbor weights were normalized inverse-distance weights. A two-layer graph head combined each bin embedding with weighted neighbor embeddings, and the full training graph was used as the input for each graph-head optimization step. In the within-group setting, each HE-ST group was divided into five spatial folds along the x-axis, and each bin was evaluated by a model trained on the remaining spatial folds from the same patient group. In the pairwise setting, the model was trained on one HE-ST group and evaluated on the other group, with the reciprocal direction evaluated in the same way. Because the two HE-ST groups came from different patients, this pairwise setting evaluated cross-patient generalization.

#### Tumor sample identification

For sample-level prediction, each sample was represented by the set of bin embeddings extracted from the frozen pretrained backbone. A graph-gated attention classifier treated all bins from one sample as a graph, connected each bin to its eight nearest bins by Euclidean distance, and aggregated the graph into a sample-level representation. The model input was the sample graph of bin embeddings and spatial coordinates. The output was a sample-level label, such as tumor status, T stage, Crohn’s disease activity stage or a clinical marker. All sample-level evaluations used patient-level folds to avoid assigning samples from the same patient to different evaluation folds. Tumor-sample prediction was evaluated in two settings. In the standard out-of-fold setting, tumor presence was evaluated with patient-grouped five-fold cross-validation. In the clinical-mock setting, one patient fold was held out as the test set; models were trained with different train/validation splits over the remaining patient folds, and predictions for the held-out patients were averaged across the resulting models. T-stage prediction used tumor samples and patient-grouped five-fold cross-validation. Crohn’s disease activity-stage prediction used Crohn’s disease samples and patient-grouped five-fold cross-validation. Recurrence or metastasis and lymphovascular-invasion prediction used tumor samples with patient-grouped three-fold cross-validation.

### Interpretability of AlphaFISH

#### Feature ablation

To estimate how individual gene or microbe features affected model output, a model-level feature-ablation analysis was performed. For a trained model, the predicted tumor probability *p*(*x*) was first computed for each bin *x*. An ablated input *x^(–f)^* was then constructed by replacing all occurrences of a feature *f*, either a gene or a microbe, with a zero vector representing feature absence. All other features were left unchanged. The model-derived contribution score was defined as

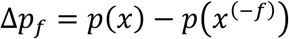

where a positive Δ*p_f_* indicates that the presence of *f* increases the predicted tumor probability, and a negative Δ*p_f_* suggests a protective or nontumor-associated effect. At the dataset level, we summarized the importance of each feature by aggregating its effects across quasi-single-cells and slides. Specifically, for the feature *f* we computed Δ*p_f_* across all quasi-single-cells/slides in which it appears, and then averaged this score over the five cross-validation models to obtain a stable importance estimate. Features were ranked according to *mean*(Δ*p_f_*) − *std*(Δ*p_f_*) for downstream interpretation.

#### Attention-based interaction analysis

To characterize differences in interaction patterns between tumor and non-tumor cells, we performed an attention-based interaction analysis. Specifically, from the last transformer layer, we extracted raw self-attention weights, which captured intra-cell interactions among features within a quasi-single-cell, and raw cross-attention weights, which captured inter-cell interactions between a center cell and its niche cells. For each attention head, we applied row-and column-wise rank normalization to make scores comparable across positions, and then averaged the normalized attention maps across heads. We then compared tumor and non-tumor cells by examining the resulting attention scores for gene–gene, gene–microbe, and microbe–microbe pairs, and summarized tumor vs. non-tumor differences by aggregating these pairwise statistics across the five seeded models.

### Bioinformatics analysis

Differential gene expression analysis (related to **Figure 4-6**) was conducted using the Seurat^42^ package. The *FindAllMarkers* function from Seurat was used to identify differentially expressed genes between clusters. This function uses a nonparametric Wilcoxon rank-sum test to assess differences in gene expression between groups. The analysis results were then filtered to identify genes with a log-fold change greater than 1 and a p-value less than 0.05, indicating substantial and statistically significant expression differences.

For analysis of Figure 2, Scanpy^43^ in Python was used to analyze mRNA expression and bacterial abundance data derived from over 10,000,000 nanoprobes (binned into 670,000 quasi-single-cell spots, namely qSC-spots, via 4×4 binning) obtained from 149 sampling trials across 68 subjects. The UMAP (Uniform Manifold Approximation and Projection) algorithm was applied to visualize mRNA expression and bacterial abundance data by embedding samples into a low-dimensional space, revealing sample clustering based on transcriptional expression. Unsupervised clustering analysis of the mRNA expression data from the 670,000 qSC-spots was also performed using *Scanpy*. Based on the multidimensional space formed by the 41 host genes and 51 bacterial genera, the Scanpy clustering algorithm was used to process the mRNA expression data. To visualize and compare these clustering patterns, the UMAP embedding method was used to reduce the dimensionality of the mRNA expression data, enabling clearer observation and comparison of distribution differences across clusters in a two-dimensional space.

### Cell type classification

To characterize the cellular composition of the spatial multi-omics dataset, we constructed a supervised cell-type classifier utilizing a comprehensive public single-cell RNA-seq dataset^27^ as the reference atlas. Feature selection was restricted to the intersection of genes shared between the reference atlas and the target spatial dataset. To ensure comparability across platforms, gene expression values were standardized using Z-score normalization. We implemented a Random Forest classifier to learn cell-type-specific signatures from the reference data. The model was trained and validated using a stratified splitting strategy (80% training, 20% validation) to ensure robust performance across all cell lineages. The classifier achieved an overall accuracy of 82% on the independent validation set (**Suppl. Figure S13**). Furthermore, performance was assessed using normalized confusion matrices to verify high sensitivity and specificity for each cell type. The validated classifier was subsequently applied to our spatial multi-omics dataset to predict each cell’s identity, serving as the foundation for downstream spatial analysis.

### Statistics and producibility

At least three independent biological replicates were used for all experiments. Statistical significance for comparing different experimental conditions was determined using the appropriate hypothesis test, as indicated in the Figure captions. A p-value less than 0.05 was considered statistically significant; all p-values are indicated in the respective Figures.

## Acknowledgement

This work was supported by General Research Fund (11215920, 11220024, 11218522, 11218523) from the Research Grants Council of Hong Kong SAR, and support from Innovation and Technology Commission of Hong Kong through the Centre for Cerebro-Cardiovascular Health Engineering and funds from City University of Hong Kong (7005084, 7005206, 7005642, 7020003, 7020077, 9680233, 9240060) are also acknowledged.

## Author contribution

P.S. conceived the project and supervised the research. F.G., H.S., and P.S. designed experiments. F.G., H.S., and C.H. carried out the investigation and analyzed the data. C.H., M.H., and J.M. developed the computational model. Y.W., X.H., C.X., F.G., and X.D. contributed to different aspects of the experimental works. J.K. provided the clinical resources. H.S., F.G., and P.S. wrote the manuscript. C.H. J.M. and L.Y.M. contributed to the writing of the manuscript.

## Code availability

Code for AlphaFISH is available via the following anonymous link: https://anonymous.4open.science/r/AlphaFISH_paper-B558/README.md

## Data availability

The data that support the findings of this study are available from the corresponding author on request.

