## Supplementary material for "Cohort-scale Spatial Host-Microbiome Predicts Post-Resection Recurrence in Colorectal Cancer": Supplemeatary

Guo *et al.*

Correspondence should be addressed to
Jianzhu Ma
Peng Shi

**Supplementary figures**

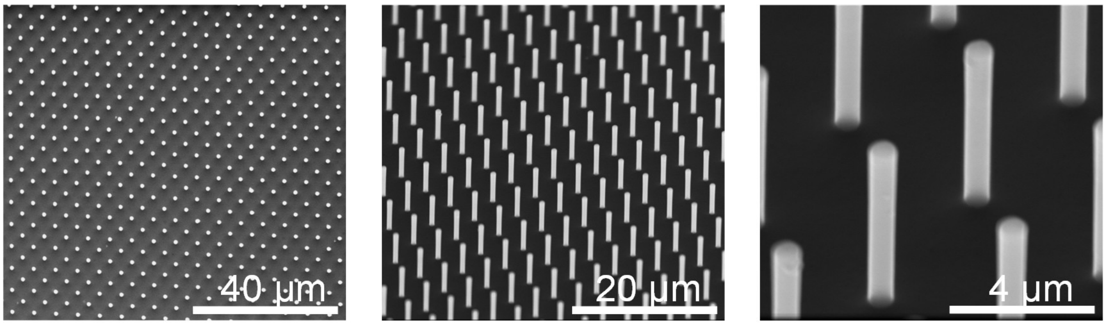


**Supplementary Figure S1.** SEM characterization of the nanostructured biochip. Each nanoprobe has a diameter of 0.8 µm and is arranged with a uniform 4 µm pitch, enabling high-resolution capture of spatially resolved molecular information. Scale bars: 40 µm (left), 20 µm (middle), and 4 µm (right).

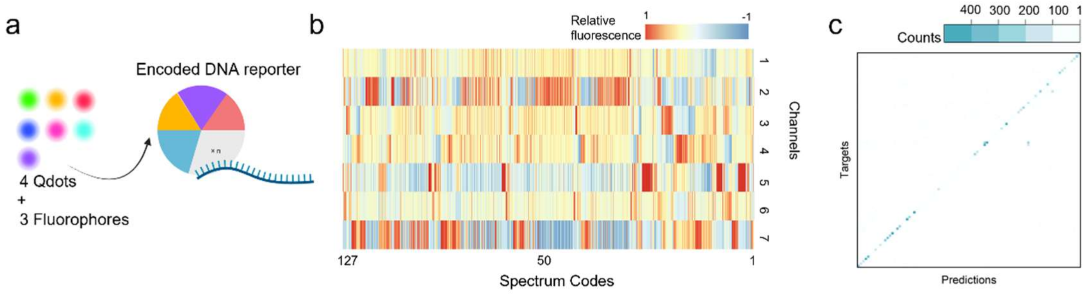


**Supplementary Figure S2.** SpectrumCODE multiplexing strategy in the AlphaFISH. **(a)** Schematic of the combinatorial encoding strategy, where 4 quantum dots and 3 organic fluorophores are utilized to construct encoded DNA reporters. **(b)** Heatmap illustrating the spectral profiles of the SpectrumCODE library in AlphaFISH. The heatmap displays normalized relative fluorescence intensities across seven confocal imaging channels for multiple replicates of each of the 127 unique SpectrumCODE. Each column corresponds to an individual measurement (replicate) of a specific spectrum code, with rows representing the seven spectral channels. Distinct barcode identities are grouped and ordered along the x-axis, highlighting spectral separability and reproducibility across replicates. (**c**) Classification performance of the SpectrumCODE decoding model. Scatter plot comparing the ground-truth number of targets (x-axis) with the machine learning predicted counts (y-axis). Strong linear correlation is observed (diagonal dashed line), indicating high decoding accuracy across a wide dynamic range.

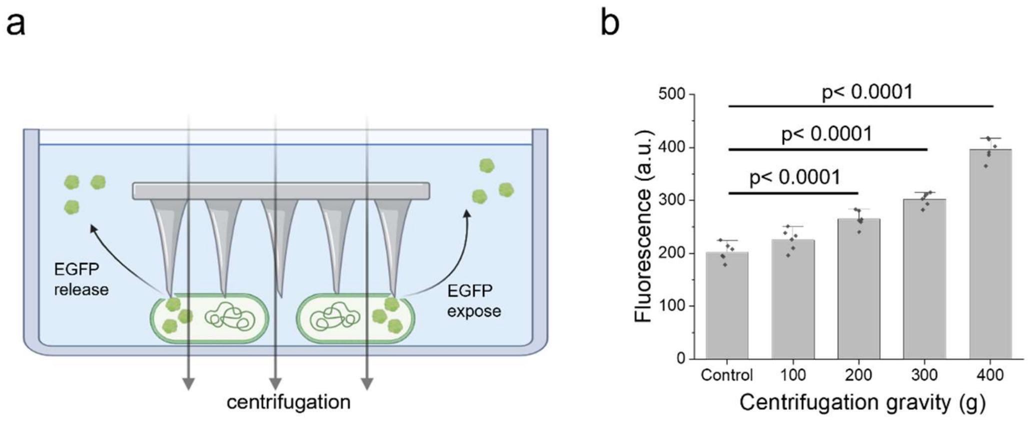


**Supplementary Figure S3**. Release of intracellular bacterial content induced by the biochip under centrifugation. **(a)** Schematic illustration of the mechanical exposing process: *E. coli* expressing EGFP are shown releasing their intracellular content upon penetration by the nanoprobe array during centrifugation. **(b)** Quantification of exposing efficiency. Fluorescence intensity measurements demonstrate that significant release of intracellular content (indicated by EGFP leakage) is achieved at centrifugal forces >200g, confirming the biochip's capacity for efficient extraction of bacterial RNAs. Data are presented as mean±SD, analyzed by two-tailed t-test, n=6.

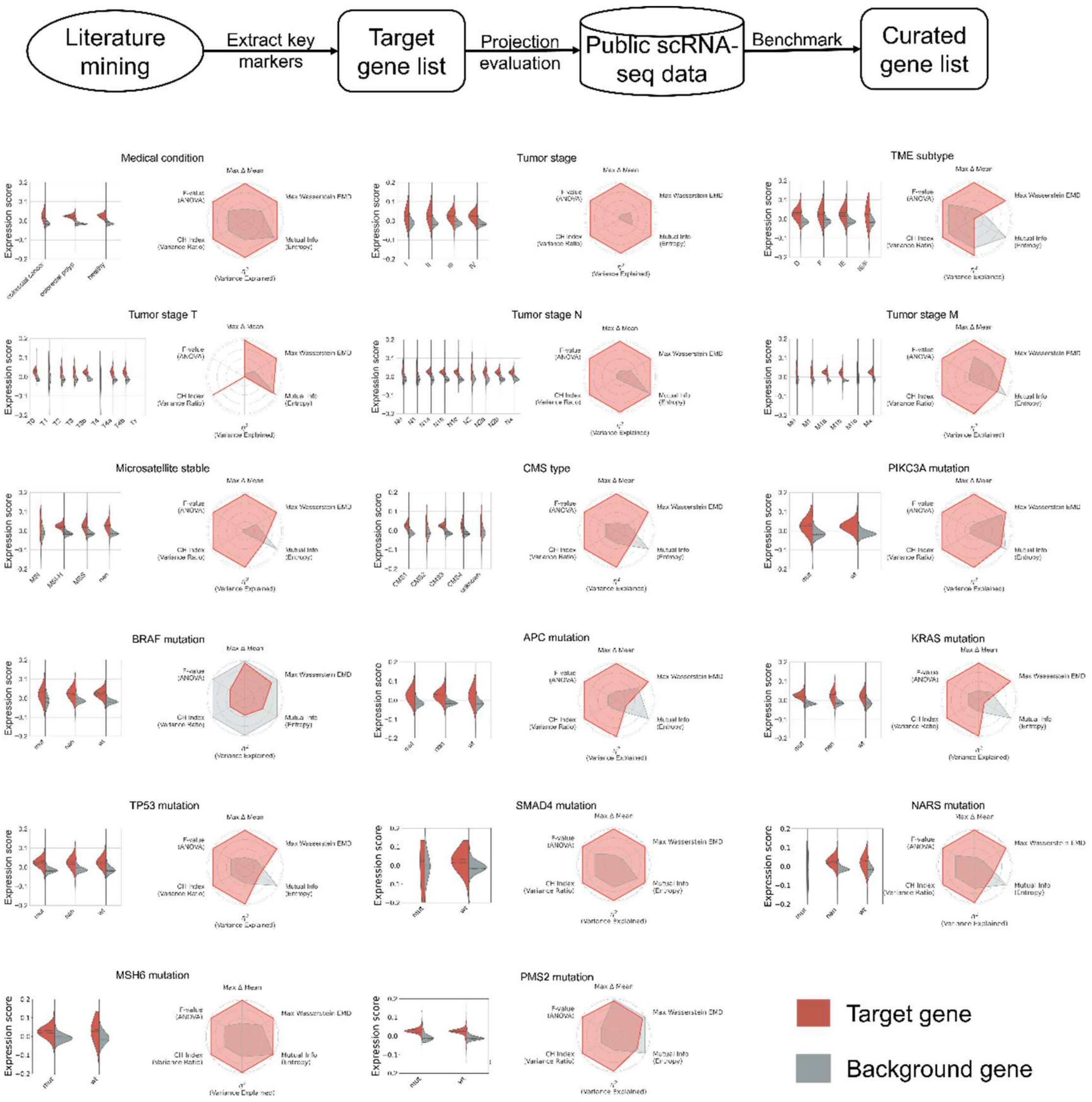


**Supplementary Figure S4.** 41-gene module consistently exhibits significantly higher activity scores compared to matched background gene sets across diverse clinical subgroups, including various medical conditions (cancer, polyp, healthy), tumor stages, molecular subtypes (e.g., CMS, MSI status), and major cell types (all pairwise comparisons, Mann-Whitney U or Kruskal-Wallis tests, $p < 0.001). This uniform upregulation—as opposed to stochastic or heterogeneous variations—demonstrates that the module captures a biologically coherent signal robustly induced in disease contexts. Radar plots confirm that the module's discriminative power significantly exceeds that of random background sets, yielding higher effect sizes (e.g., Cohen's d, AUC, and mutual information) across all categorical labels examined (e.g., TNM stage, KRAS mutation status). Together, these results validate the module as a sensitive and specific molecular signature for patient stratification, prognosis, and the resolution of cell-type-specific transcriptional states.

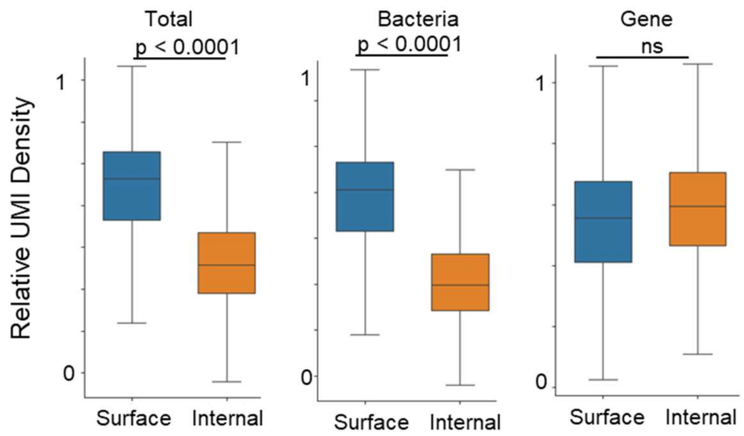


**Supplementary Figure S5.** Assessment of spatial bias in UMI capture between tissue surface and internal regions. Relative UMI density comparison between the tissue surface and internal regions for total reads, bacterial RNA, and host gene transcripts. The surface-associated microbiome exhibits significantly higher UMI counts compared to the internal tissue ($p < 0.0001$, Wilcoxon rank-sum test), whereas no significant difference (ns) is observed in the distribution of host gene transcripts. This pattern confirms that AlphaFISH's touch-and-go profiling effectively captures native surface-associated bacterial populations, minimizing the spatial bias and microbial loss typically encountered during conventional tissue processing. Each box indicates the first quartile, median, and third quartile of the data; the whisker limit is 1.5 times the interquartile range.

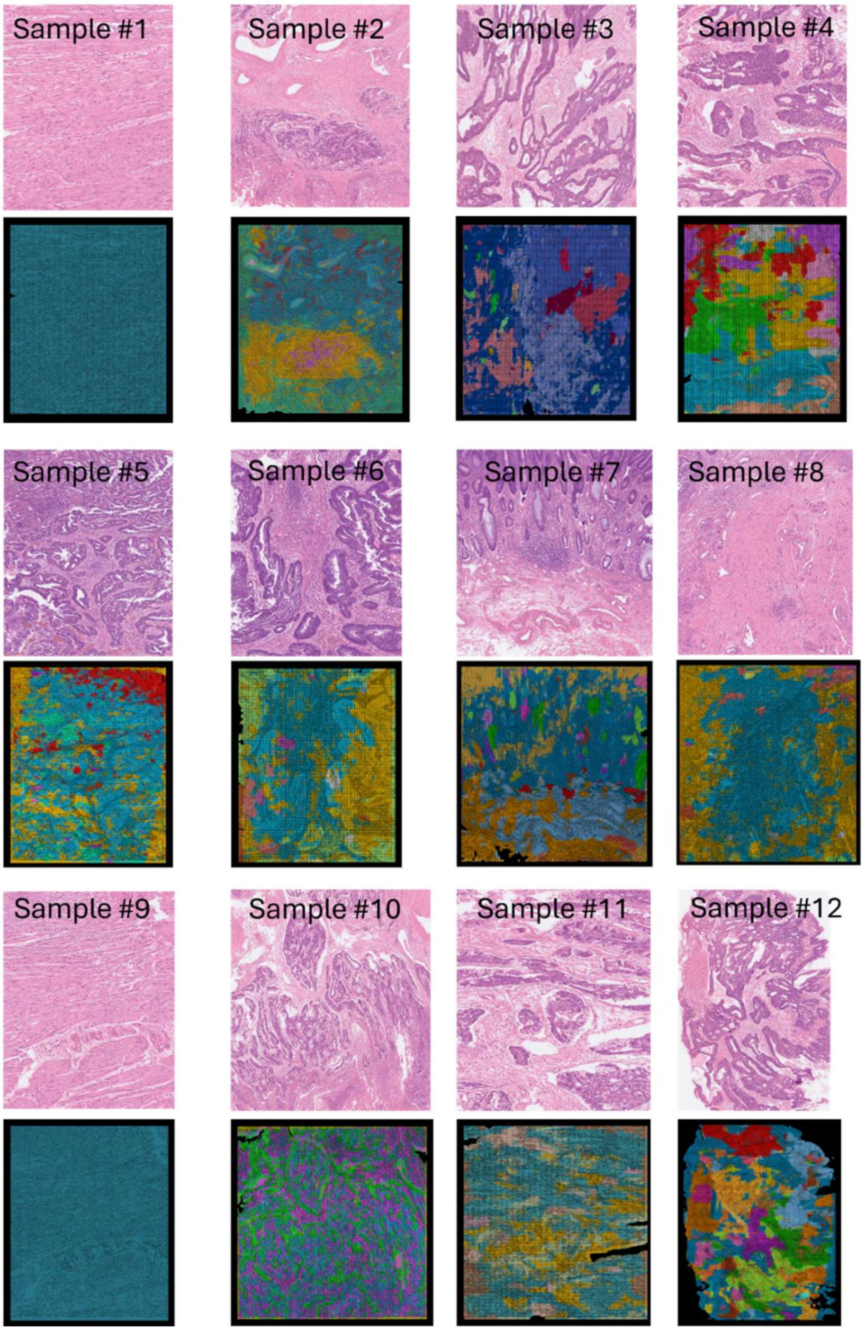


**Supplementary Figure S6.** Spatial alignment of AlphaFISH dual-omics maps with H&E-stained histological features. High-resolution spatial transcriptomic (STrp) and microbial (SMib) signal maps generated by AlphaFISH (lower panels) demonstrate robust concordance with corresponding H&E-stained tissue sections (upper panels).

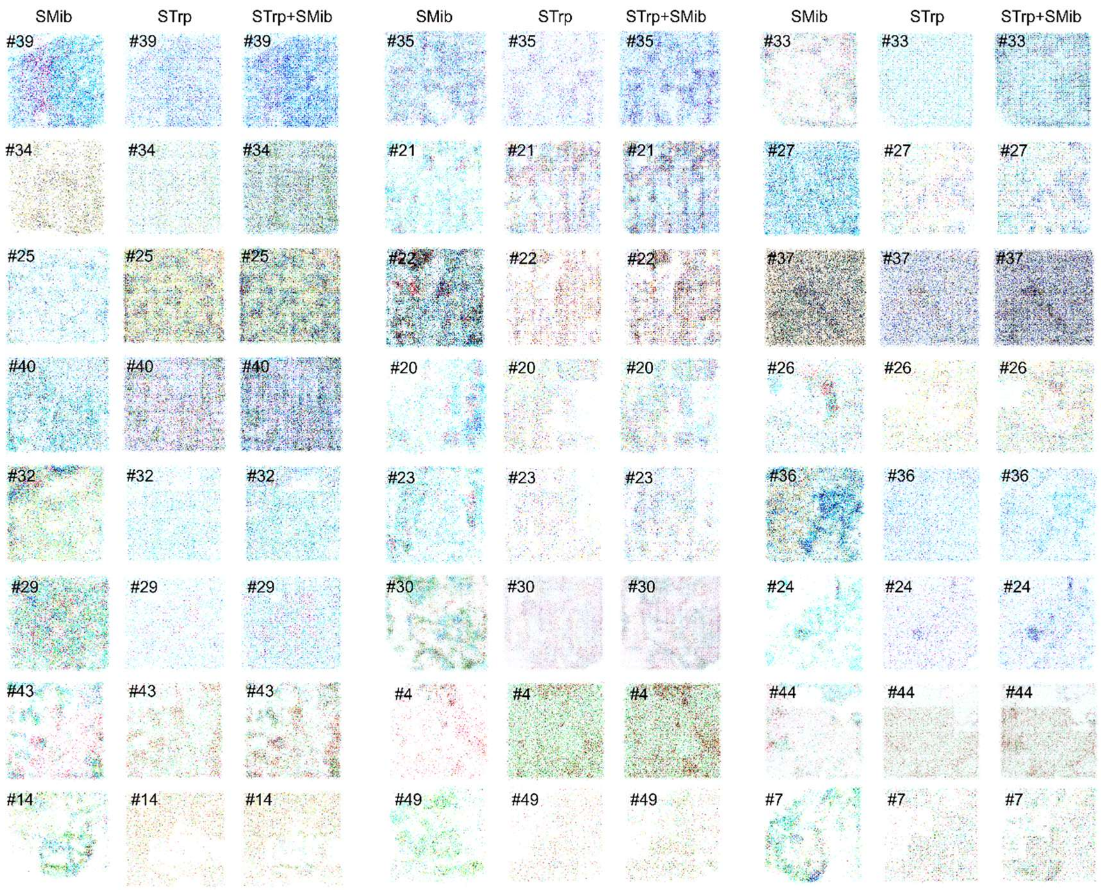


**Supplementary Figure S7.** More examples of dual-omics spatial profiling of transcriptomics and microbiome in CRC patient biopsies, showing the spatial maps of raw transcriptomic (STrp) and microbiome (SMib) signals**.** Patient identifiers (#) are annotated in the top-left corner of each sample.

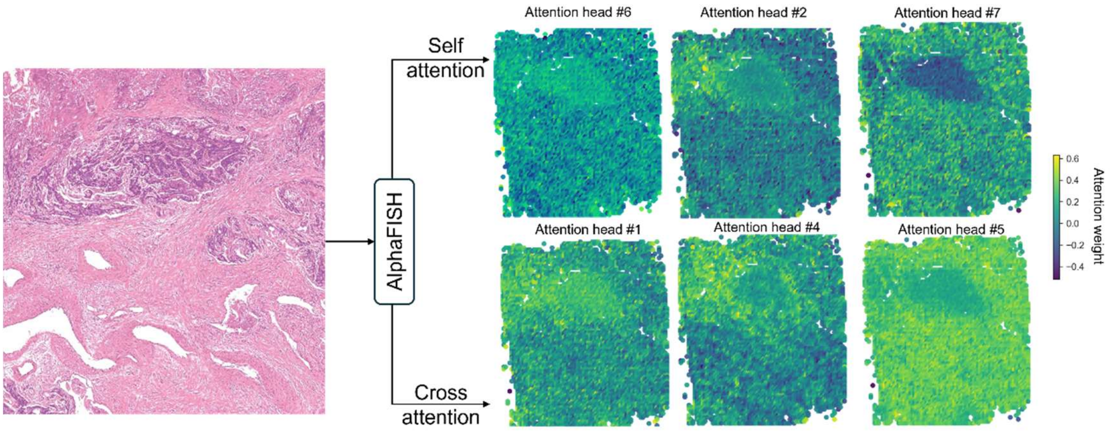


**Supplementary Figure S8. Attention heatmaps of AlphaFISH.** The visualization displays representative attention weights from the model's multi-head mechanism. While the majority of the 12 attention heads preferentially focus on tumor regions, specific heads demonstrate specialized spatial attention. In the cross-attention module, head #4 highlights tumor-adjacent tissues, whereas head #5 focuses on non-tumor areas. Similarly, within the self-attention module, head #2 targets tumor-adjacent regions, and head #7 attends to non-tumor areas, indicating the model's capacity to disentangle complex tissue heterogeneity.

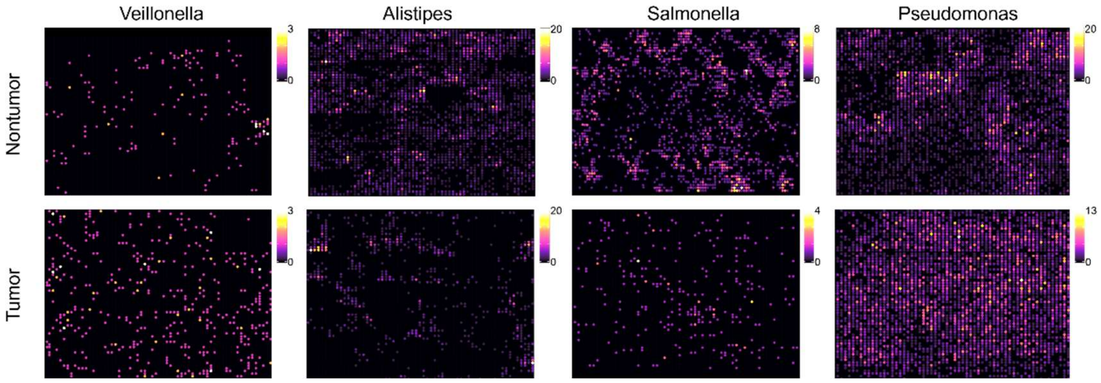

**Supplementary Figure S9.** Representative spatial expression (color indexed) of top-ranked (from **Figure 4**) bacteria features, in paired Tumor or Nontumor samples.

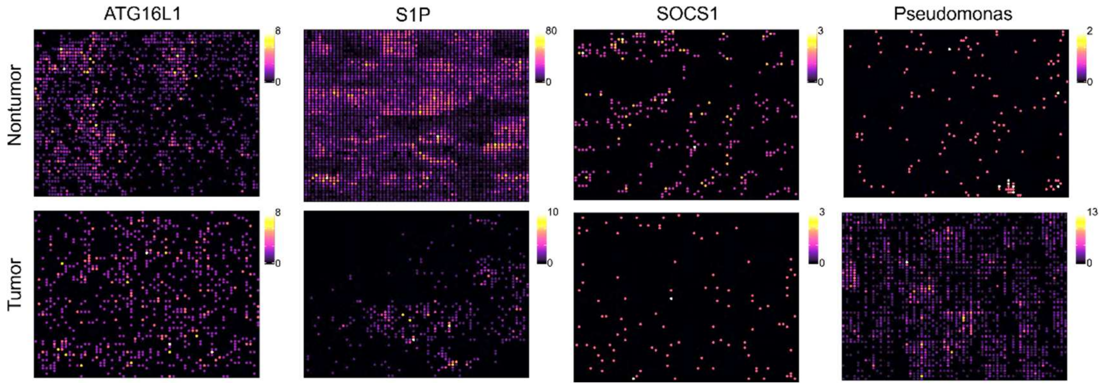

**Supplementary Figure S10.** Representative spatial expression (color indexed) of top-ranked (from **Figure 4**) gene features, in paired Tumor or Nontumor samples.

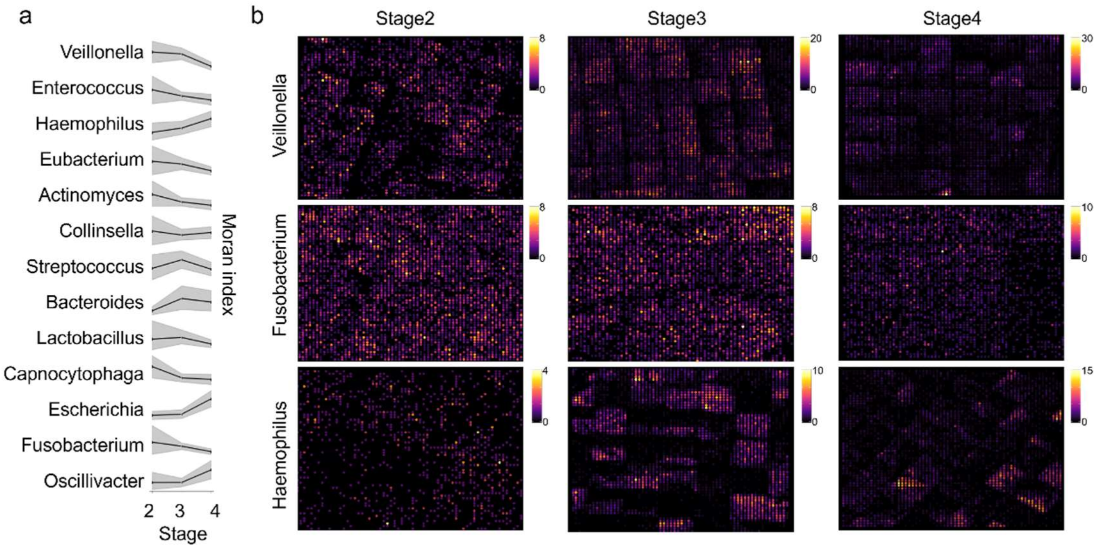


**Supplementary Figure S11.** Stage-dependent evolution of bacteria distribution. **(a)** Trend analysis illustrating the changing Moran index of top-ranked bacteria (from **Figure 5**) across tumor stages 2, 3, and 4. Shaded areas represent confidence intervals. **(b)** Representative spatial expression (color index) of bacteria across different clinical stages.

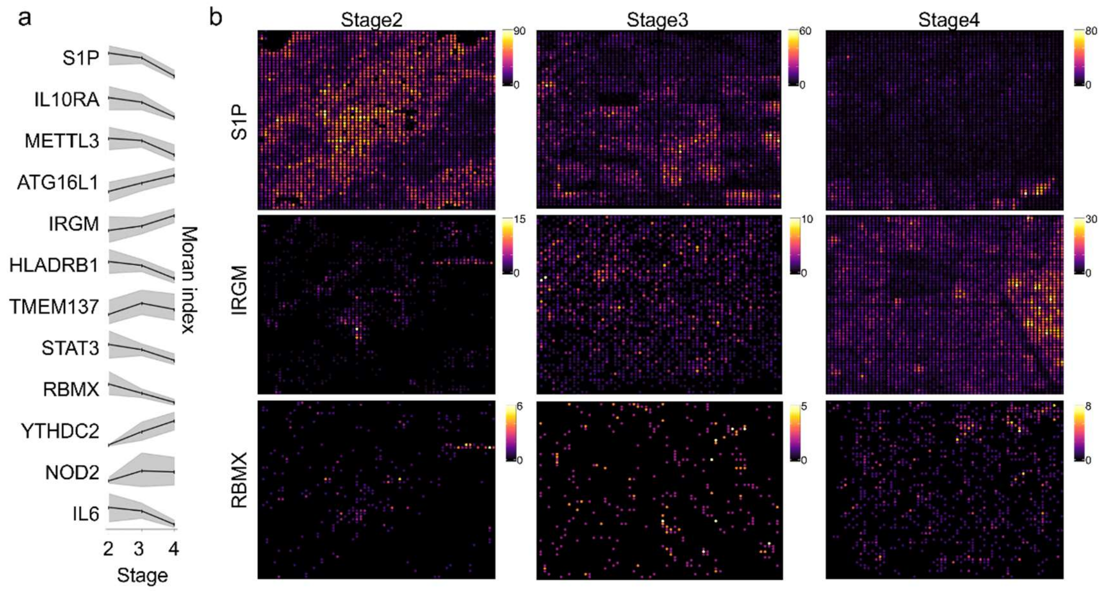


**Supplementary Figure S12.** Stage-dependent evolution of gene distribution. **(a)** Trend analysis illustrating the changes in the Moran index for top-ranked genes (from **Figure 5**) across clinical stages 2, 3, and 4. Shaded regions indicate confidence intervals. **(b)** Representative spatial expression (color index) of genes across different clinical stages.

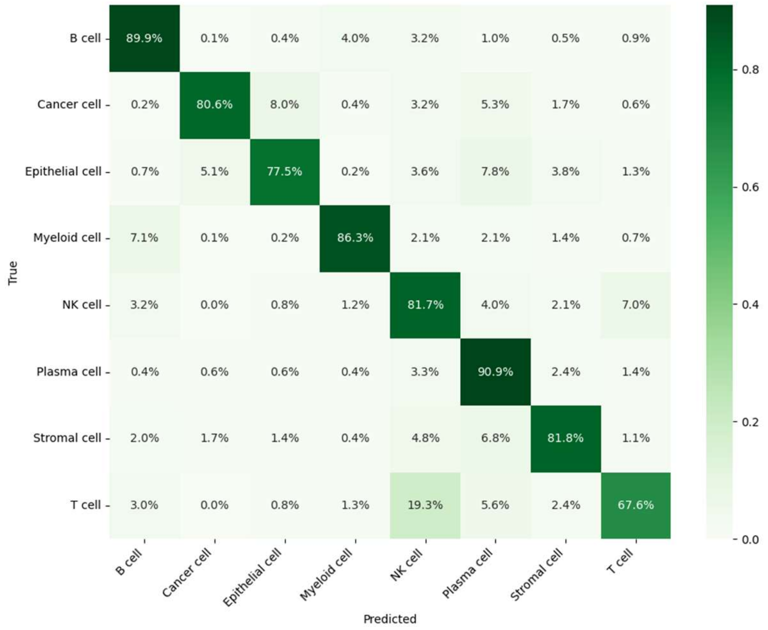


**Supplementary Figure S13.** Performance evaluation of the cell-type classification model. The confusion matrix illustrates the classification accuracy of the Random Forest classifier on the independent validation set (20% stratified split). The Y-axis represents the ground truth (True) cell identity, while the X-axis represents the model-predicted identity.

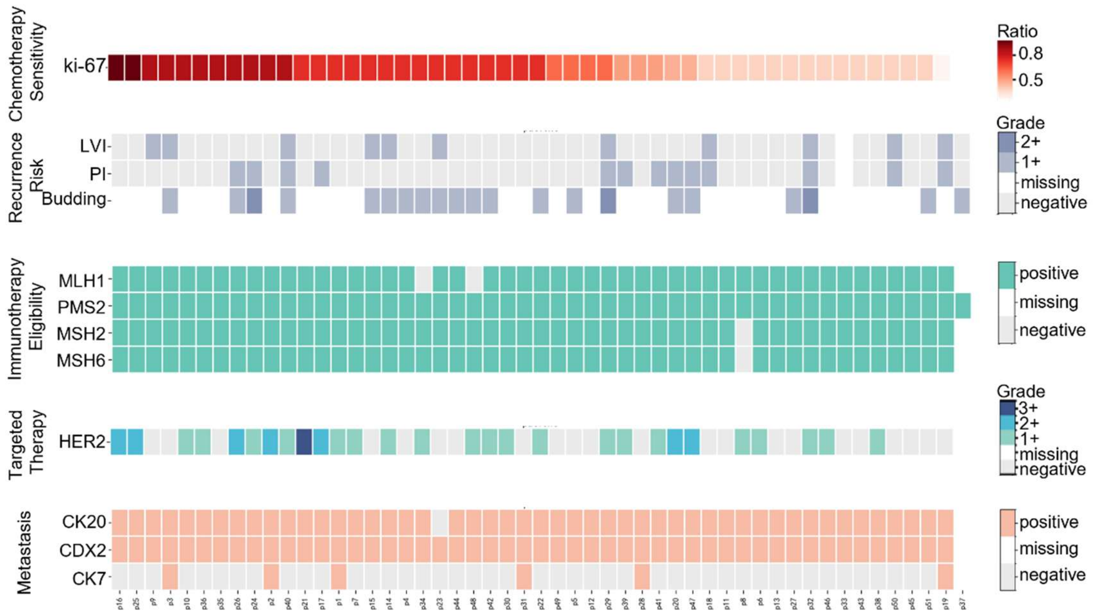


**Supplementary Figure S14.** Comprehensive clinic pathological landscape of the CRC patient cohort. The heatmap details a comprehensive panel of standard clinical and immunohistochemical (IHC) biomarkers across individual patients (columns). To contextualize the baseline clinical baseline, these features are stratified into five primary functional domains driving conventional treatment decisions: (1) Chemotherapy Sensitivity, indicated by the Ki-67 proliferation index; (2) Recurrence Risk, encompassing lymphovascular invasion (LVI), perineural invasion (PI), and tumor budding; (3) Immunotherapy Eligibility, assessed via mismatch repair (MMR) protein proficiency (MLH1, PMS2, MSH2, and MSH6); (4) Targeted Therapy, evaluated by HER2 expression grading; and (5) Metastasis and Tumor Differentiation, indicated by cytokeratin and transcription factor profiles (CK20, CDX2, and CK7). Distinct color scales denote the continuous expression ratio (for Ki-67), histological severity grading (e.g., 1+ to 3+ for HER2 and risk factors), or binary clinical status (positive/negative for MMR and metastasis markers).

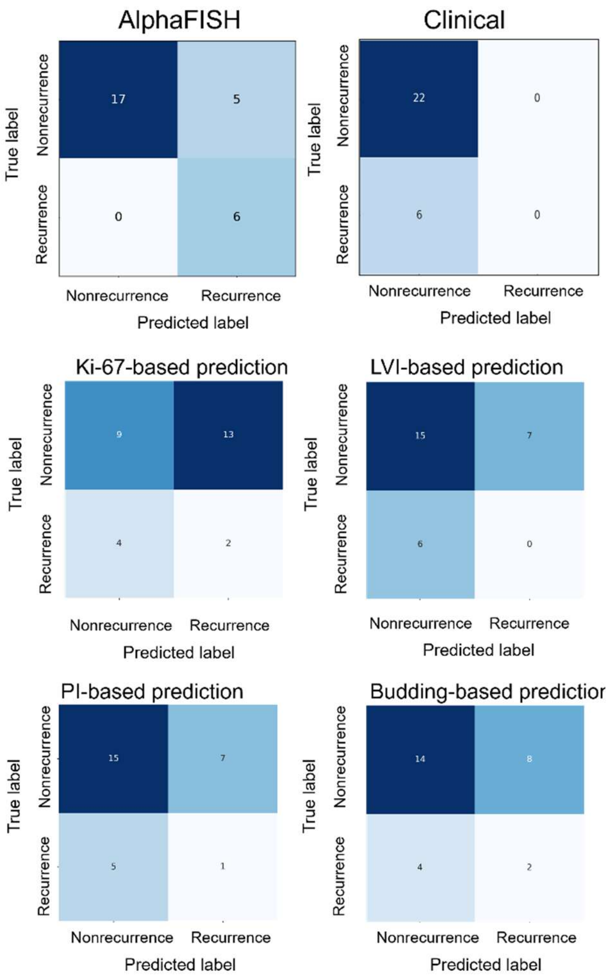


**Supplementary Figure S15**. Performance comparison of tumor recurrence prediction between AlphaFISH and conventional clinico-pathological metrics. Confusion matrices evaluating the predictive accuracy of AlphaFISH against standard clinical indicators. (**a-b)** Predictive performance of the AlphaFISH model (**a**) contrasted with a composite Clinical model integrating Ki-67, lymphovascular invasion (LVI), pathological index (PI), and tumor budding (**b**). (**c–f)** Independent predictive performance of individual clinical metrics: Ki-67 proliferation index (**c**), LVI (**d**), PI (**e**), and tumor budding (**f**). Each matrix illustrates the distribution of true versus predicted patient outcomes (Nonrecurrence vs. Recurrence).

### Supplementary tables

**Supplementary Table S1.** List of gene encoding and associated reporter sequences.

| Gene target | Encoded Reporter Sequence (5' to 3') | Spectrum-code |
|---|---|---|
| APOA1 | GACACGCACTTCCGCACATTCCGTTCGCACGCCGATTACC | 100 |
| ATG16L1 | CAGATAGATAGTCCACTGAATGGGAAGGTGACGAATGAGG | 101 |
| CBLL1 | GACAATGAGTTACAAGGCACTAATAGTTCTGGATCCTTGGG | 110 |
| DUOX2 | AGAAGGAAGCAGCCAAAGATGGAGTGCCAGCGATGGAGTG | 111 |
| DUOXA2 | CATGCCGCAGGCTTCAGCGTTCCACTGCTCATCGTTATTC | 1000 |
| E-cadherin | CAGGTGGGATGAGGATGAGCAGTGGTCGGTGGTGGTGGAG | 1001 |
| FMR1 | CGGGGCTCCAATGGCGCTTTCTACAAGGCATTTGTA | 1010 |
| Foxo3a | TCTACAGCAGCTCAGCCAGCCTGTCACCTTCAGTAAGCAA | 1011 |
| FTO | CGCGGAGGAACGAGAGCGGGAAGCTAAGAAACTGAG | 1100 |
| GPR84 | CCAAGCTCCGTACCCGATTCAACCTGCTCATAGCCAACC | 1101 |
| HLA-DRB1 | CAGCACGTACCTCTATAACCGCCAGCGGAAGATCAAGAAA | 1110 |
| HNRNPA2B1 | ACCACAGAAGAAAGTTTGAGAAACTACTATGAGCAATGGGG | 1111 |
| IGF2BP1 | ATGAACAAGCTTTACATCGGCAACCTCAACGAGAGT | 10000 |
| IGF2BP2 | ATGATGAACAAGCTGTACATTGGGAACCTGAGCCCC | 10001 |
| IGF2BP3 | ACTGAATGGATTCCAGTTAGAGAACTTCACCTTGAAGGTTG | 10010 |
| IL10RA | CCTCCACTGGACACCCATCCCAAATCAGTCTGAAAGTACC | 10011 |
| IL10RB | CTTGACGGAATGTGATTTCTCAAGTCTTTCCAAGTATGGT | 10100 |
| IL23R | TTCCTGTGAAATGAGATACAAGGCTACAACAAACCAAACT | 10101 |
| IL6 | GTTGCCTTCTCCCTGGGGCTGCTCCTGGTGTTGCCTGCTG | 10110 |
| IRGM | GTCTGCCACCACAACCCTGGAGAACTACCTGATGGAAATG | 10111 |
| MAdCAM1 | CCACGCAGGGAGAAGTGATCCCAACAGGCTCGTCCAAA | 11000 |
| METTL14 | AATAGCAAAGATGAACAGAGGGAGATTGCTGAAACCAGA | 11001 |
| METTL16 | TCAATGCATGCAAGAAATAGATACAAGGACAAACCACCTGA | 11010 |
| METTL3 | ATGTCGGACACGTGGAGCTCTATCCAGGCCCATAAG | 11011 |
| NOD2 | ACCTCAATGACGATGCGGACACTGTGCTGGTGGTGGGTGA | 11100 |
| PRRC2A | GGGCCGACTGCCAAGGGGAAGGATGGAAAGAAGTAT | 11101 |
| PTPN22 | GCTGGCTGTGGAAGGACTGGTGTTATTTGTGCTATTGATT | 11110 |
| RBM15B | GAGGACCTGATGCCGGAGGATGATCAGAGAGCCACT | 11111 |
| RBMX | GGTGGGCTTAATACAGAGACGAATGAGAAAGCCCTT | 100000 |
| S1P | GTATGAACTGAGCAATGTGCAAGAAGATAGCCAACCAATGTG | 100001 |
| Smad7 | CGGATCTGAAGGCGCTCACGCACTCGGTGCTCAAGAAA | 100010 |
| SMOX | GCATCAGCCTCTATTCCAAGAATGGCGTGGCCTGCTACCT | 100011 |
| SOCS1 | CGACACGCACTTCCGCACATTCCGTTCGCACGCCGATTAC | 100100 |
| SREBP2 | AAGTCCTTCAGCCTCAAGTCCAAAGCCTGGTGACATCCTC | 100101 |
| STAT3 | ACGTCATTAGCAGAATCTCAACTTCAGACCCGTCAACAAA | 100110 |
| TMEM137 | GAGGAGCTGGCAGCCCTCTTTGCGCCCTACGGCACGGTCA | 100111 |
| TTC7A | AGAAGACCACAAATAACAGCACGTCGAGGCATCTGAAAGG | 101000 |
| VCAM-1 | TATCTTGCTCAGATTGGTGACTCCGTCTCATTGACTTGCA | 101001 |
| VEGF-A | TCCAGGAGTACCCTGATGAGATCGAGTACATCTTCAAGCC | 101010 |
| YTHDC2 | CTGGAGCGCTTCCGCTACGGTGACCAGAGAGAAATG | 101011 |
| YTHDF1 | AATGACTTTGAGCCCTACCTTTCTGGACAGTCCAAT | 101100 |

**Supplementary Table S2.** Annotations of all mRNA transcripts profiled in this study.

| mRNA | Function group | Functional relevance to colon disease |
|---|---|---|
| YTHDF1 | m6A RNA methylation reader | stem cell homeostasis, intestinal barrier integrity [1] |
| YTHDF2 | | intestinal epithelial cell proliferation and apoptosis [2, 3] |
| YTHDF3 | | mRNA translation and degradation [1, 3] |
| YTHDC1 | | inflammatory responses, epithelial barrier integrity [4] |
| YTHDC2 | | RNA metabolism [5, 6] |
| RBMX | | RNA splicing and metabolic regulation [5, 7] |
| HNRNPA2B1 | | macrophage polarization, mRNA stability of pro-inflammatory cytokines [7, 8] |
| IGF2BP1 | | maintains intestinal barrier function by stabilizing occluding mRNA [9] |
| IGF2BP2 | | macrophage conversion, ferroptosis [6, 7, 10] |
| IGF2BP3 | | RNA stability and translation [10, 11] |
| FMR1 | | m6A regulation [6, 7, 12] |
| PRRC2A | | epithelial cell regeneration and repair [6, 13] |
| METTL3 | m6A RNA methylation writer | inflammatory response, signaling transduction [6, 12, 14] |
| METTL14 | | stem cell apoptosis, mucosal barrier function [6, 12] |
| METTL16 | | m6A modification of U6 snRNA and mRNAs [6, 7] |
| WTAP | | key component of the m6A writer complex [6, 12, 15] |
| VIRMA | | m6A modification, inflammation-related gene expression [1, 6, 7] |
| ZC3H13 | | intestinal epithelial cell regeneration and repair [1, 6, 7] |
| CBLL1 | | immune cell signaling, cytokine production [3, 6, 7] |
| RBM15 | | inflammation regulation, immune response [1, 6, 7] |
| RBM15B | | intestinal homeostasis, inflammatory responses [1, 6, 7] |
| FTO | m6A RNA methylation eraser | immune response, signaling transduction and autophagy [1, 7, 12] |
| MAdCAM1 | inflammation regulation | lymphocyte recruitment to the intestinal mucosa [16, 17] |
| XIAP | | cytokines, signaling transduction, apoptosis [18-20] |
| GPR84 | | pro-inflammatory responses in immune cells [21-23] |
| Smad7 | | immune response, cytokines, signaling transduction [24, 25] |
| Foxo3a | | inflammatory responses in monocytes, cytokines [26, 27] |
| TMEM229B | | cellular signaling, membrane transport, or cell-cell interactions [28] |
| IL10RA | | immune response, signal transduction, cytokines [29-31] |
| IL10RB | | immune response, signal transduction, cytokines [31, 32] |
| IL23R | | JAK-STAT signaling, pro-inflammatory cytokine production [33] |
| IL6 | | pro-inflammatory, T cell activation [34] |
| STAT3 | | adaptive immunity, cytokines [35] |
| SOCS1 | | immune tolerance in the intestine [36] |
| APOA1 | | anti-immune response [37] |
| DUOXA2 | | thyroid hormone synthesis, immune regulation [38] |
| DUOX2 | | reactive oxygen species in the intestine [38] |
| TMEM137 | | cytokine production, innate immune response [39] |
| TTC7A | cell proliferation | cell differentiation, immune response [40] |
| SMOX | | microbiome homeostasis [41] |
| ICAM-1 | | cell proliferation, apoptosis, signal transduction [42] |
| SREBP2 | | signaling transduction [43] |
| VCAM-1 | | cytokines, signal transduction, cell adhesion [44] |
| E-cadherin | | integrity of the intestinal barrier [45] |
| VEGF-A | | cell adhesion, cytokines, immune response [46] |
| S1P | | cytokines, signaling transduction [47] |
| ATG16L1 | | anti-bacterial autophagic response, cytokines [48] |
| HLA-DRB1 | | adaptive immunity [49] |
| INAVA | | cytokine, signaling transduction [50] |
| NOD2 | | Immune regulation, susceptibility to inflammation [19, 30] |
| PTPN22 | | cytokine production, immune cell activation [30] |

**Supplementary Table S3.** List of bacteria encoding.

| Genus | Family | Order | Class | Phylum | Spectrum -code |
|---|---|---|---|---|---|
| Acidaminococcus | Acidaminococcaceae | Selenomonadales | Negativicutes | Firmicutes | 101101 |
| Actinomyces | Actinomycetaceae | Actinomycetales | Actinomycetes | Actinomycetota | 101110 |
| Akkermansia | Akkermansiaceae | Verrucomicrobiales | Verrucomicrobiae | Verrucomicrobiota | 101111 |
| Alistipes | Rikenellaceae | Bacteroidales | Bacteroidia | Bacteroidota | 110000 |
| Alloprevotella | Prevotellaceae | Bacteroidales | Bacteroidia | Bacteroidota | 110001 |
| Anaerotruncus | Oscillospiraceae | Eubacteriales | Clostridia | Firmicutes | 110010 |
| Atopobium | Atopobiaceae | Coriobacteriales | Coriobacteriia | Actinobacteria | 110011 |
| Bacteroides | Bacteroidaceae | Bacteroidales | Bacteroidia | Bacteroidota | 110100 |
| Barnesiella | Barnesiellaceae | Bacteroidales | Bacteroidia | Bacteroidota | 110101 |
| Bifidobacterium | Bifidobacteriaceae | Bifidobacteriales | Actinobacteria | Actinobacteria | 110110 |
| Bilophila | Desulfovibrionaceae | Desulfovibrionales | Desulfovibrionia | Thermodesulfobacteriota | 110111 |
| Butyrivibrio | Lachnospiraceae | Clostridiales | Clostridia | Firmicutes | 111000 |
| Capnocytophaga | Flavobacteriaceae | Flavobacteriales | Flavobacteriia | Bacteroidota | 111001 |
| Cardiobacterium | Cardiobacteriaceae | Cardiobacteriales | Gammaproteobacteria | Pseudomonadota | 111010 |
| Chlamydia | Chlamydiaceae | Chlamydiales | Chlamydiia | Chlamydiae | 111011 |
| Clostridium | Clostridiaceae | Clostridiales | Clostridia | Firmicutes | 111100 |
| Collinsella | Coriobacteriaceae | Coriobacteriales | Coriobacteriia | Actinomycetota | 111101 |
| Coprobacter | Barnesiellaceae | Bacteroidales | Bacteroidia | Bacteroidota | 111110 |
| Corynebacterium | Corynebacteriaceae | Corynebacteriales | Actinobacteria | Actinobacteria | 111111 |
| Desulfovibrio | Desulfovibrionaceae | Desulfovibrionales | Desulfovibrionia | Thermodesulfobacteriota | 1000000 |
| Dialister | Veillonellaceae | Veillonellales | Negativicutes | Bacillota | 1000001 |
| Enterococcus | Enterococcaceae | Lactobacillales | Bacilli | Firmicutes | 1000010 |
| Escherichia | Enterobacteriaceae | Enterobacteriales | Gammaproteobacteria | Proteobacteria | 1000011 |
| Eubacterium | Eubacteriaceae | Clostridiales | Clostridia | Firmicutes | 1000100 |
| Faecalibacterium | Ruminococcaceae | Clostridiales | Clostridia | Firmicutes | 1000101 |
| Fibrobacter | Fibrobacteraceae | Fibrobacterales | Fibrobacteria | Fibrobacteres | 1000110 |
| Fusobacterium | Fusobacteriaceae | Fusobacteriales | Fusobacteriia | Fusobacteria | 1000111 |
| Haemophilus | Pasteurellaceae | Pasteurellales | Gammaproteobacteria | Proteobacteria | 1001000 |
| Helicobacter | Helicobacteraceae | Campylobacterales | Epsilonproteobacteria | Campylobacterota | 1001001 |
| Kingella | Neisseriaceae | Neisseriales | Betaproteobacteria | Pseudomonadota | 1001010 |
| Lactobacillus | Lactobacillaceae | Lactobacillales | Bacilli | Firmicutes | 1001011 |
| LIS | Lachnospiraceae | Lachnospirales | Clostridia | Bacillota | 1001100 |
| Marvinbryantia | Lachnospiraceae | Lachnospirales | Clostridia | Bacillota | 1001101 |
| Mycobacterium | Mycobacteriaceae | Corynebacteriales | Actinobacteria | Actinobacteria | 1001110 |
| Neisseria | Neisseriaceae | Neisseriales | Betaproteobacteria | Pseudomonadota | 1001111 |
| Oscillibacter | Oscillospiraceae | Eubacteriales | Clostridia | Bacillota | 1010000 |
| Prevotella | Prevotellaceae | Bacteroidales | Bacteroidia | Bacteroidota | 1010001 |
| Propionibacterium | Propionibacteriaceae | Propionibacteriales | Actinomycetes | Actinomycetota | 1010010 |
| Pseudobutyrivibrio | Lachnospiraceae | Clostridiales | Clostridia | Firmicutes | 1010011 |
| Pseudomonas | Pseudomonadaceae | Pseudomonadales | Gammaproteobacteria | Pseudomonadota | 1010100 |
| Rikenella | Rikenellaceae | Bacteroidales | Bacteroidia | Bacteroidota | 1010101 |

**Supplementary Table S4.** Detailed Clinical Characteristics of the CRC Cohort.

| Subject No. | Sample counts | Tumor location | Age | T | M | N |
|---|---|---|---|---|---|---|
| 1 | 2 | ascending colon | 70-74 | 4 | 1 | 0 |
| 2 | 2 | rectum | 46-50 | 4 | 2 | |
| 3 | 2 | rectum | 70-74 | 4 | 1 | 0 |
| 4 | 2 | sigmoid colon | 51-55 | 4 | 1 | |
| 5 | 2 | sigmoid colon | 61-65 | 4 | 1 | 0 |
| 6 | 2 | rectum | 80-84 | 4 | 2 | 0 |
| 7 | 2 | descending colon | 51-55 | 4 | 0.5 | 0 |
| 8 | 2 | descending colon | 66-70 | 4 | 2 | 0 |
| 9 | 2 | transverse colon | 80-84 | 4 | 0 | 1 |
| 10 | 2 | ascending colon | 71-75 | 4 | 2 | |
| 11 | 2 | rectum | 41-45 | 4 | 2 | |
| 12 | 2 | rectum | 66-70 | 4 | 2 | |
| 13 | 2 | ascending colon | 61-65 | 4 | 2 | |
| 14 | 2 | transverse colon | 71-75 | 4 | 2 | |
| 15 | 2 | descending colon | 51-55 | 4 | 1 | |
| 16 | 2 | sigmoid colon | 66-70 | 3.5 | 2 | 0 |
| 17 | 2 | descending colon | 71-75 | 3.5 | 2 | |
| 18 | 2 | descending colon | 71-75 | 3.5 | 2 | 0 |
| 19 | 2 | ascending colon | 66-70 | 3.5 | 0 | |
| 20 | 2 | sigmoid colon | 76-80 | 3 | 2 | 0 |
| 21 | 2 | ascending colon | 21-25 | 3 | 2 | |
| 22 | 2 | rectum | 71-75 | 3 | 2 | 0 |
| 23 | 2 | sigmoid colon | 61-65 | 3 | 0 | |
| 24 | 2 | sigmoid colon | 66-70 | 3 | 0 | |
| 25 | 2 | rectum | 66-70 | 3 | 0 | |
| 26 | 2 | rectum | 66-70 | 3 | 1 | |
| 27 | 2 | rectum | 71-75 | 3 | 0 | |
| 28 | 2 | sigmoid colon | 56-60 | 3 | 1 | |
| 29 | 2 | sigmoid colon | 61-65 | 3 | 1 | |
| 30 | 2 | sigmoid colon | 41-45 | 3 | 0 | 0 |
| 31 | 2 | rectum | 71-75 | 3 | 1 | 0 |
| 32 | 2 | sigmoid colon | 66-70 | 3 | 0 | 0 |
| 33 | 2 | sigmoid colon | 61-65 | 3 | 1 | 0 |
| 34 | 2 | sigmoid colon | 46-50 | 3 | 1 | |
| 35 | 2 | rectum | 66-70 | 3 | 1 | 0 |
| 36 | 2 | sigmoid colon | 91-95 | 3 | 1 | |
| 37 | 2 | rectum | 51-55 | 3 | 0 | 0 |
| 38 | 2 | rectum | 76-80 | 3 | 1 | 0 |
| 39 | 2 | rectum | 71-75 | 3 | 0 | |
| 40 | 2 | sigmoid colon | 56-60 | 3 | 1 | |
| 41 | 2 | rectum | 66-70 | 3 | 1 | |
| 42 | 2 | ascending colon | 71-75 | 3 | 2 | |
| 43 | 2 | rectum | 56-60 | 3 | 1 | |
| 44 | 2 | descending colon | 51-55 | 2.5 | 0 | |
| 45 | 2 | ascending colon | 56-60 | 2.5 | 1 | |
| 46 | 2 | rectum | 71-75 | 2 | 0 | 0 |
| 47 | 2 | rectum | 51-55 | 2 | 0 | |
| 48 | 2 | ascending colon | 41-45 | 2 | 0 | |
| 49 | 2 | descending colon | 46-50 | 2 | 1 | |
| 50 | 2 | sigmoid colon | 71-75 | 1.5 | 0 | 0 |
| 51 | 2 | rectum | 66-70 | 1.5 | 0 | |

Note: TNM staging was determined based on preoperative imaging (CT/MRI/PET-CT) and assigned by experienced clinicians according to the AJCC 8th edition criteria. Staging reflects clinical stage (cTNM) prior to surgical or pathological confirmation.

**Supplementary Table S5.** Summary for subjects in CD cohort.

| Subject No. | Sample No. | Age | Sampling location | CD Status | SES-CD # |
|---|---|---|---|---|---|
| 1 | 1 | 41-45 | stomach | active | 4 |
| 1 | 2 | 41-45 | ileum | active | 4 |
| 2 | 3 | 31-35 | ileum | active | 3 |
| 2 | 4 | 31-35 | ascending colon | active | 4 |
| 3 | 5 | 41-45 | ascending colon | active | 4 |
| 3 | 6 | 41-45 | rectum | active | 4 |
| 4 | 7 | 31-35 | sigmoid colon | remission | 2 |
| 5 | 8 | 26-30 | ascending colon | remission | 1 |
| 6 | 9 | 31-35 | stomach | active | 3 |
| 6 | 10 | 31-35 | ileum | active | 4 |
| 6 | 11 | 31-35 | ileum | active | 4 |
| 6 | 12 | 31-35 | descending colon | active | 3 |
| 7 | 13 | 41-45 | ileum | active | 4 |
| 7 | 14 | 41-45 | ileum | active | 4 |
| 7 | 15 | 41-45 | transverse colon | active | 3 |
| 7 | 16 | 41-45 | sigmoid colon | active | 4 |
| 8 | 17 | 46-50 | ileum | active | 3 |
| 8 | 18 | 46-50 | ascending colon | active | 4 |
| 8 | 19 | 46-50 | rectum | active | 4 |
| 9 | 20 | 21-25 | cecum | active | 3 |
| 9 | 21 | 21-25 | ascending colon | active | 3 |
| 9 | 22 | 21-25 | transverse colon | active | 4 |
| 9 | 23 | 21-25 | rectum | active | 4 |
| 10 | 24 | 21-25 | stomach | active | 3 |
| 10 | 25 | 21-25 | ileum | active | 4 |
| 10 | 26 | 21-25 | ascending colon | active | 4 |
| 10 | 27 | 21-25 | transverse colon | active | 4 |
| 10 | 28 | 21-25 | sigmoid colon | active | 3 |
| 11 | 29 | 26-30 | ileum | active | 4 |
| 11 | 30 | 26-30 | ileum | active | 4 |
| 11 | 31 | 26-30 | ileum | active | 4 |
| 12 | 32 | 31-35 | ascending colon | healthy | 0 |
| 12 | 33 | 31-35 | ascending colon | healthy | 0 |
| 13 | 34 | 41-45 | ascending colon | healthy | 0 |
| 13 | 35 | 41-45 | ascending colon | healthy | 0 |
| 14 | 36 | 26-30 | ileum | remission | 2 |
| 14 | 37 | 26-30 | transverse colon | remission | 1 |
| 15 | 38 | 51-55 | stomach | remission | 1 |
| 15 | 39 | 51-55 | ileum | remission | 2 |
| 15 | 40 | 51-55 | transverse colon | remission | 1 |
| 15 | 41 | 51-55 | rectum | remission | 1 |
| 16 | 42 | 16-20 | ileum | active | 3 |
| 16 | 43 | 16-20 | ascending colon | active | 4 |
| 16 | 44 | 16-20 | transverse colon | active | 4 |
| 16 | 45 | 16-20 | sigmoid colon | active | 4 |
| 17 | 46 | 21-25 | ileum | active | 3 |
| 17 | 47 | 21-25 | ascending colon | active | 3 |

#As part of clinical care, patients were assessed using the Simple Endoscopic Score for Crohn's Disease (SES-CD), evaluated across all segments. Subjects with an SES-CD $\geq$ 3 were classified as active, while those with an SES-CD $\leq$ 2 were considered in remission. Healthy subjects exhibited an SES-CD of 0.

**Supplementary Table S6.** Clinic pathology characteristics of CRC patients with follow-up data

| Patient | Sample | Tissue | Age | Sex | Ki-67 | LVI | PI | Budding | Recurrence |
|---|---|---|---|---|---|---|---|---|---|
| p1 | n86 | Tumor | 71-75 | M | 0.7 | Neg | Neg | NA | Yes |
| p2 | n39 | Tumor | 76-80 | M | 0.8 | Neg | Neg | NA | No |
| p5 | n92 | Tumor | 71-75 | M | 0.6 | Neg | Neg | G1 | Yes |
| p6 | n95 | Tumor | 51-55 | F | 0.4 | Neg | Neg | NA | Yes |
| p7 | n79 | Tumor | 61-65 | M | 0.7 | Neg | Neg | NA | No |
| p8 | n70 | Tumor | 80-84 | F | 0.4 | Neg | Neg | NA | No |
| p9 | n68 | Tumor | 66-70 | M | 0.8 | Pos | Neg | NA | No |
| p10 | n97 | Tumor | 71-75 | M | 0.8 | Neg | Neg | NA | Yes |
| p11 | n98 | Tumor | 71-75 | F | 0.4 | Neg | Neg | NA | No |
| p12 | n67 | Tumor | 51-55 | F | 0.6 | Neg | Neg | NA | No |
| p14 | n74 | Tumor | 71-75 | M | 0.7 | Pos | Neg | G1 | No |
| p16 | n81 | Tumor | 66-70 | M | 0.9 | Neg | Neg | NA | No |
| p17 | n102 | Tumor | 66-70 | F | 0.7 | Neg | Pos | NA | No |
| p19 | n16 | Tumor | 71-75 | M | 0.3 | Pos | Pos | NA | No |
| p20 | n20 | Tumor | 66-70 | M | 0.5 | Neg | Pos | G1 | Yes |
| p22 | n17 | Tumor | 56-60 | M | 0.7 | Neg | Neg | G1 | No |
| p23 | n23 | Tumor | 61-65 | F | 0.7 | Pos | Neg | G1 | No |
| p24 | n10 | Tumor | 41-45 | M | 0.8 | Neg | Pos | G2 | No |
| p27 | n13 | Tumor | 51-55 | F | 0.4 | Neg | Neg | G1 | No |
| p28 | n14 | Tumor | 66-70 | M | 0.5 | Neg | Neg | NA | Yes |
| p29 | n26 | Tumor | 61-65 | M | 0.6 | Pos | Pos | G2 | No |
| p30 | n29 | Tumor | 66-70 | F | 0.7 | Neg | Neg | NA | No |
| p31 | n12 | Tumor | 46-50 | F | 0.7 | Neg | Neg | NA | No |
| p32 | n22 | Tumor | 66-70 | M | 0.4 | Pos | Pos | G2 | No |
| p35 | n5 | Tumor | 56-60 | M | 0.8 | Neg | Neg | NA | No |
| p38 | n2 | Tumor | 71-75 | M | 0.4 | Neg | Neg | NA | No |
| p39 | n4 | Tumor | 41-45 | F | 0.5 | Neg | Pos | NA | No |
| p40 | n19 | Tumor | 51-55 | M | 0.8 | Pos | Pos | G1 | No |

CRC, colorectal cancer; Neg, negative; Pos, positive; NA, not assessed; G1/G2, budding grade 1/2.

**Supplementary Table S7.** Prognostic significance of key pathological markers

| Abbreviation | Full Name | Definition / Clinical Significance | Reference(s) |
|---|---|---|---|
| Ki-67 | Ki-67 Proliferation Index | Nuclear antigen expressed in proliferating cells (G1, S, G2, M phases); the labeling index reflects tumor cell proliferation rate. Higher Ki-67 expression is associated with aggressive tumor behavior and serves as an independent prognostic marker in colorectal cancer. | 51 |
| LVI | Lymphovascular Invasion | Presence of tumor cells within the lymphatic or blood vessel lumina; an independent adverse prognostic indicator for lymph node metastasis and recurrence in colorectal cancer. | 52 |
| PI | Perineural Invasion | Tumor cell infiltration along nerves or within the perineural space; associated with increased local recurrence and reduced disease-free survival in colorectal cancer. | 53 |
| Budding | Tumor Budding | Isolated single cells or small cell clusters (<5 cells) at the invasive tumor front; reflects epithelial-mesenchymal transition and is a strong predictor of lymph node metastasis and poor prognosis. Graded per ITBCC guidelines: G1 (low), G2 (intermediate), G3 (high). | 54 |

**Supplementary Table S8.** Model Hyperparameters and Architecture Details

| Category | Parameter | Value | Description |
|---|---|---|---|
| Model Architecture | Embedding size | 768 | Dimension of token and hidden embeddings |
| | Hidden size | 3072 | Feed-forward network intermediate dimension |
| | Number of attention heads | 12 | Number of parallel attention heads per layer |
| | Number of Transformer layers | 6 | Stack depth of Transformer blocks |
| | Cell embedding style | max-pool | Pooling method for aggregating token embeddings into cell-level representation |
| Training | Learning rate | $10^{-4}$ | Initial/base learning rate for AdamW optimizer |
| | Number of epochs | 50 | Total training epochs |
| | Number of workers | 16 | DataLoader workers for parallel data loading |
| | Mask ratio | 0.2 | Proportion of values masked during self-supervised pretraining |
